# SARS-CoV-2 Variant Vaccine Boosters Trial: Preliminary Analyses

**DOI:** 10.1101/2022.07.12.22277336

**Authors:** Angela R. Branche, Nadine G. Rouphael, David J. Diemert, Ann R. Falsey, Cecilia Losada, Lindsey R. Baden, Sharon E. Frey, Jennifer A. Whitaker, Susan J. Little, Evan J. Anderson, Emmanuel B. Walter, Richard M. Novak, Richard Rupp, Lisa A. Jackson, Tara M. Babu, Angelica C. Kottkamp, Anne F. Luetkemeyer, Lilly C. Immergluck, Rachel M. Presti, Martín Bäcker, Patricia L. Winokur, Siham M. Mahgoub, Paul A. Goepfert, Dahlene N. Fusco, Elissa Malkin, Jeffrey M. Bethony, Edward E. Walsh, Daniel S. Graciaa, Hady Samaha, Amy C. Sherman, Stephen R. Walsh, Getahun Abate, Zacharoula Oikonomopoulou, Hana M. El Sahly, Thomas C.S. Martin, Christina A. Rostad, Michael J. Smith, Benjamin G. Ladner, Laura Porterfield, Maya Dunstan, Anna Wald, Tamia Davis, Robert L. Atmar, Mark J. Mulligan, Kirsten E. Lyke, Christine M. Posavad, Megan A. Meagher, David S. Stephens, Kathleen M. Neuzil, Kuleni Abebe, Heather Hill, Jim Albert, Teri C. Lewis, Lisa A. Giebeig, Amanda Eaton, Antonia Netzl, Samuel H. Wilks, Sina Türeli, Mamodikoe Makhene, Sonja Crandon, Marina Lee, Seema U. Nayak, David C. Montefiori, Mat Makowski, Derek J. Smith, Paul C. Roberts, John H. Beigel, the COVAIL Study Group

## Abstract

**Background:** Protection from SARS-CoV-2 vaccines wanes over time and is compounded by emerging variants including Omicron subvariants. This study evaluated safety and immunogenicity of SARS-CoV-2 variant vaccines.

**Methods:** This phase 2 open-label, randomized trial enrolled healthy adults previously vaccinated with a SARS-CoV-2 primary series and a single boost. Eligible participants were randomized to one of six Moderna COVID19 mRNA vaccine arms (50µg dose): Prototype (mRNA-1273), Omicron BA.1+Beta (1 or 2 doses), Omicron BA.1+Delta, Omicron BA.1 monovalent, and Omicron BA.1+Prototype. Neutralization antibody titers (ID_50_) were assessed for D614G, Delta, Beta and Omicron BA.1 variants and Omicron BA.2.12.1 and BA.4/BA.5 subvariants 15 days after vaccination.

**Results:** From March 30 to May 6, 2022, 597 participants were randomized and vaccinated. Median age was 53 years, and 20% had a prior SARS-CoV-2 infection. All vaccines were safe and well-tolerated. Day 15 geometric mean titers (GMT) against D614G were similar across arms and ages, and higher with prior infection. For uninfected participants, Day 15 Omicron BA.1 GMTs were similar across Omicron-containing vaccine arms (3724-4561) and higher than Prototype (1,997 [95%CI:1,482-2,692]). The Omicron BA.1 monovalent and Omicron BA.1+Prototype vaccines induced a geometric mean ratio (GMR) to Prototype for Omicron BA.1 of 2.03 (97.5%CI:1.37-3.00) and 1.56 (97.5%CI:1.06-2.31), respectively. A subset of samples from uninfected participants in four arms were also tested in a different laboratory at Day 15 for neutralizing antibody titers to D614G and Omicron subvariants BA.1, BA.2.12.2 and BA.4/BA.5. Omicron <u>BA.4/BA.5 GMTs</u> were approximately one third <u>BA.1 GMTs</u> (Prototype 517 [95%CI:324-826] vs. 1503 [95%CI:949-2381]; Omicron BA.1+Beta 628 [95%CI:367-1,074] vs. 2125 [95%CI:1139-3965]; Omicron BA.1+Delta 765 [95%CI:443-1,322] vs. 2242 [95%CI:1218-4128] and Omicron BA.1+Prototype 635 [95%CI:447-903] vs. 1972 [95%CI:1337-2907).

**Conclusions:** Higher Omicron BA.1 titers were observed with Omicron-containing vaccines compared to Prototype vaccine and titers against Omicron BA.4/BA.5 were lower than against BA.1 for all candidate vaccines.

**Clinicaltrials.gov:** NCT05289037

## BACKGROUND

Severe Acute Respiratory Syndrome Coronavirus 2 (SARS-CoV-2) has infected over 500 million people worldwide and resulted in more than 6 million deaths, including greater than 1 million deaths in the United States (US).^1,2^ The vaccines authorized for emergency use or fully approved in the US are safe and highly effective against severe disease.^3-6^ However, vaccine protection against symptomatic SARS-CoV-2 infection wanes over time. ^7,8^ Additionally, several variants of concern (VOCs) have now emerged, including B.1.351 (Beta), B.1.617.2 (Delta), B.1.1.529 (Omicron BA.1) and Omicron subvariants BA.2.12.1 and BA.4/BA.5, all with mutations in the spike protein receptor binding domain,^9-11^ leading to increased risk of breakthrough infections. While additional booster doses of current vaccines, which are directed against the spike protein of the prototype Wuhan-1 SARS-CoV-2 strain, may improve vaccine effectiveness against VOCs in the short term,^12-15^ variant-specific vaccines may be needed for improved and durable protection against known and emerging VOCs.

In the first stage of this adaptive phase 2 clinical trial, we evaluated Moderna vaccines containing mRNA encoding the spike protein of different SARS-CoV-2 strains (the prototype or variant strains, alone or in combination) to evaluate the breadth and magnitude of the early antibody response.

## METHODS

### Study Design

This phase 2 open-label, randomized, clinical trial was performed at 22 sites in the US (Supplemental Table 1). Eligible participants were healthy adults 18 years of age and older (with or without a history of prior SARS-CoV-2 infection) who had received a primary series and a single homologous or heterologous boost with an approved or emergency use authorized COVID-19 vaccine (Supplemental Table 2).^16^ The most recent vaccine dose, and/or prior infection must have occurred at least 16 weeks prior to randomization. Full eligibility criteria are described at clinicaltrials.gov (NCT 05289037).

The trial was reviewed and approved by a central institutional review board and overseen by an independent Data and Safety Monitoring Board. Participants provided written informed consent before undergoing trial-related activities. The trial was sponsored and funded by the National Institutes of Health (NIH). The NIAID SARS-CoV-2 Assessment of Viral Evolution (SAVE) program team was consulted to inform study arm design and variant vaccine selection.^17^

### Enrollment and Randomization

Eligible participants were stratified by age (18-64 and ≥ 65 years) and history of confirmed SARS-CoV-2 infection, and randomly assigned in 1:1:1:1:1:1 ratio to one of six regimens. Trial vaccines included variant versions of mRNA-1273, alone or in combination, at a total dose of 50 μg in the following formulations: Arm 1, Prototype (mRNA-1273) vaccine; Arms 2 and 3, Omicron BA.1 + Beta bivalent vaccine, with Arm 3 participants receiving a second dose at Day 57; Arm 4, Omicron BA.1 + Delta bivalent vaccine; Arm 5, Omicron BA.1 monovalent vaccine; and Arm 6, Omicron BA.1 + Prototype bivalent vaccine (Supplemental Table 3). All trial vaccines were provided by Moderna (Cambridge, MA).

Blood is collected for immunogenicity assessments on Day 1 (before vaccination), Days 15 and 29 after vaccination, and 3, 6, 9 and 12 months after last vaccination. Immunologic data are currently available up to the Day 15 after first vaccination. Arms 2 and 3 have been combined for this report.

Solicited injection site and systemic solicited adverse events (AEs) were collected for 7 days after vaccination and unsolicited AEs through Day 29. Serious adverse events (SAEs), new-onset chronic medical conditions (NOCMCs), adverse events of special interest (AESIs), and medically attended adverse events (MAAEs) are being collected for the duration of the trial.

Information on antigen- or PCR- confirmed symptomatic or asymptomatic SARS-CoV-2 infection at any time after randomization was collected.

Data cut off was June 27, 2022.

### Immunogenicity Assays

SARS-CoV-2 neutralization titers, expressed as the serum inhibitory dilution required for 50% neutralization (ID_50_), were assessed using pseudotyped lentiviruses presenting SARS-CoV-2 spike mutations for the D614G, (Wuhan-1 containing a single D614G spike mutation), B1.617.2 (Delta), B.1.351 (Beta) and B.1.1.529 (Omicron BA.1) variants, as described previously.^18,19^ A random subset of samples (20-24 per selected vaccine arms, distributed equally between age strata and sites) were analyzed for neutralization titers to the Omicron BA.2.12.1 and BA.4/BA.5 subvariants in a separate laboratory.^20^ Electrochemiluminescence immunoassays (ELECSYS) were used for the detection of anti-nucleocapsid (N) (N-ELECSYS; Elecsys Anti-SARS-CoV-2 N, Roche, Indianapolis) at baseline.^19^

### Antigenic Cartography and Antibody Landscapes

Antigenic cartography is a visualization tool used to guide the vaccine strain selection of seasonal influenza viruses^21^ and is now being used to understand the relationship among SARS-CoV-2 VOCs.^22,23^ Antigenic cartography uses antibody neutralization data to position virus variants and sera relative to each other in an n-dimensional Euclidean space as previously described.^21,22^ The antigenic map from Wilks et al.^22^ was used here as basis for the map and for antibody landscapes^24^ which contains an estimate for the position of BA.4/BA.5 as described in the supplement. For each serum-variant pair, the fold-change from the maximum titer variant in a specific serum is calculated to obtain a target distance from the serum. Serum and variant coordinates are optimized such that the sum of squared errors between Euclidean map distance and this target distance is minimized. One distance unit (square in any direction) in the map corresponds to a two-fold dilution in the neutralization assay.^22^

To visualize how the different arms modify baseline immunity, antibody landscapes for each vaccine candidate were constructed. ^22^ Antibody landscapes allow an estimate of the distribution of immunity across the antigenic space by fitting a surface to a serum’s neutralization titers against the mapped variants in a third dimension.^24^ Variants that have similar titers to the same sample are positioned close to each other, hence map distance can be understood as a measure of antigenic similarity.

### Statistical Analysis

The primary objective of this study is to evaluate the magnitude, breadth and durability of SARS-CoV-2 specific antibody titers in serum samples by estimating 95% confidence intervals (CI) for the geometric mean titer (GMT) at each timepoint when samples are collected. No formal hypothesis tests were planned. Any titer above the lower limit of detection (LLOD) is considered seropositive. 95% CI of GMTs are calculated using the Student’s t-distribution. For the purpose of analysis, participants were defined as previously infected by self-report of a confirmed positive antigen or PCR testing or the detection of anti-SARS-CoV-2 N antibodies.

ANCOVA models were used to estimate GMT ratios of variant vaccines compared to Prototype vaccine and included independent variables for vaccination arms, age (18-64 years and ≥ 65 years of age), previous infection history, and baseline titers. For modeling purposes, titers were log_10_ transformed and estimated mean differences were back transformed to generate GMT ratios between vaccination groups. Unadjusted 97.5% confidence intervals based on the t-distribution are reported.

Study endpoints and sample size determination are included in the protocol.

## RESULTS

### Study Population

From March 30 to May 6, 2022, 602 participants were randomized, 597 of whom received a study vaccine (Supplemental Figure 1 CONSORT). Demographic and baseline characteristics were similar across study arms (Table 1). Median age was 53 years (range: 18-85), 35% were ≥65 years and 53% were female. Most participants were White (82%), 8% were Black or African American and 6% were Hispanic or Latinx. The majority of participants (95%) had received an mRNA-based primary series and boost vaccine (89% were homologous). Twenty-percent were defined as previously infected by anti-N antibody seropositivity at baseline and/or by a PCR- or antigen-confirmed COVID-19 episode. Median duration (range) between study vaccination and the last prior vaccination or infection was 168 (110-333) days. Median follow up duration at data cut off is 70 days.

**TABLE 1:**
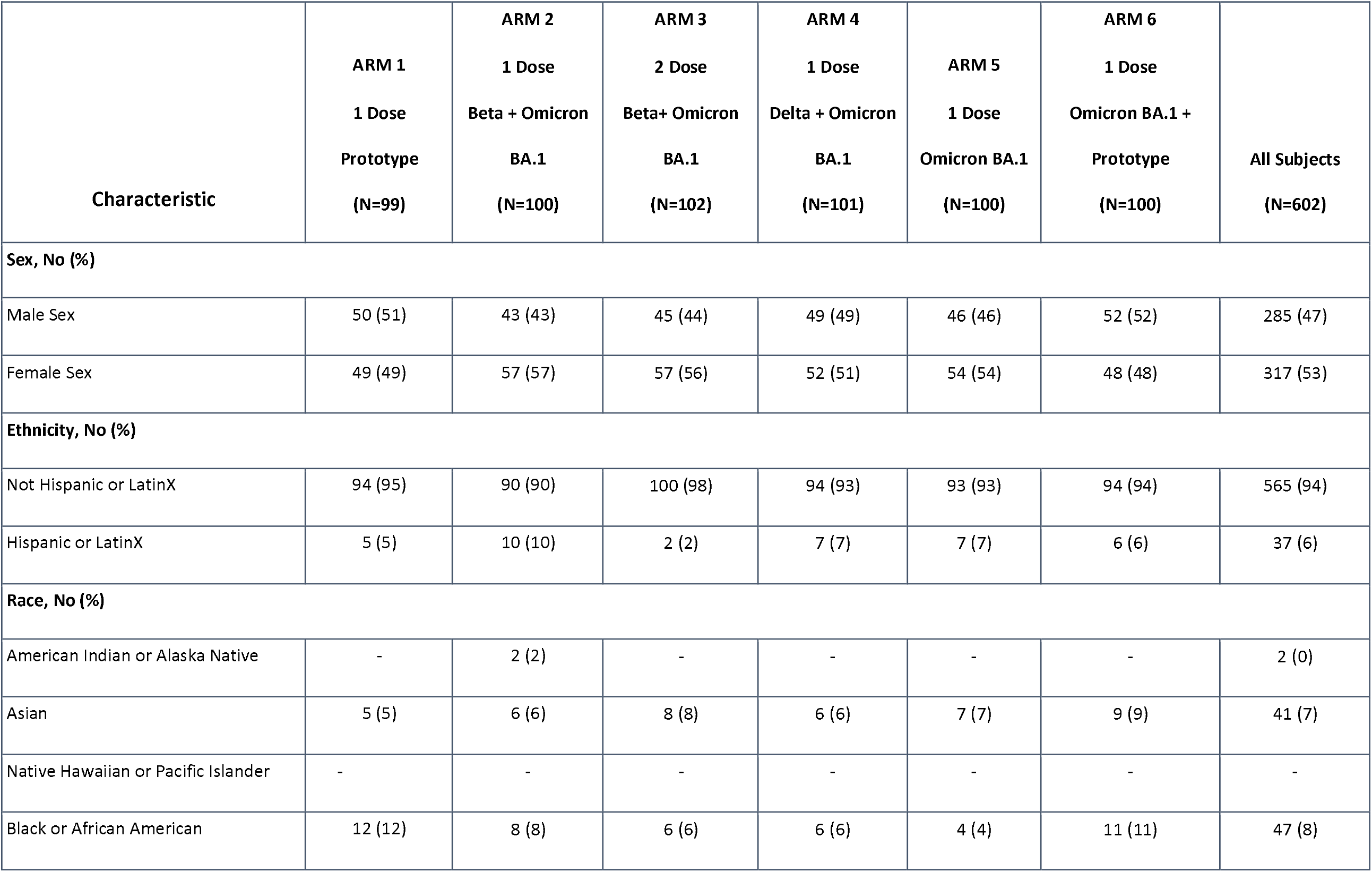
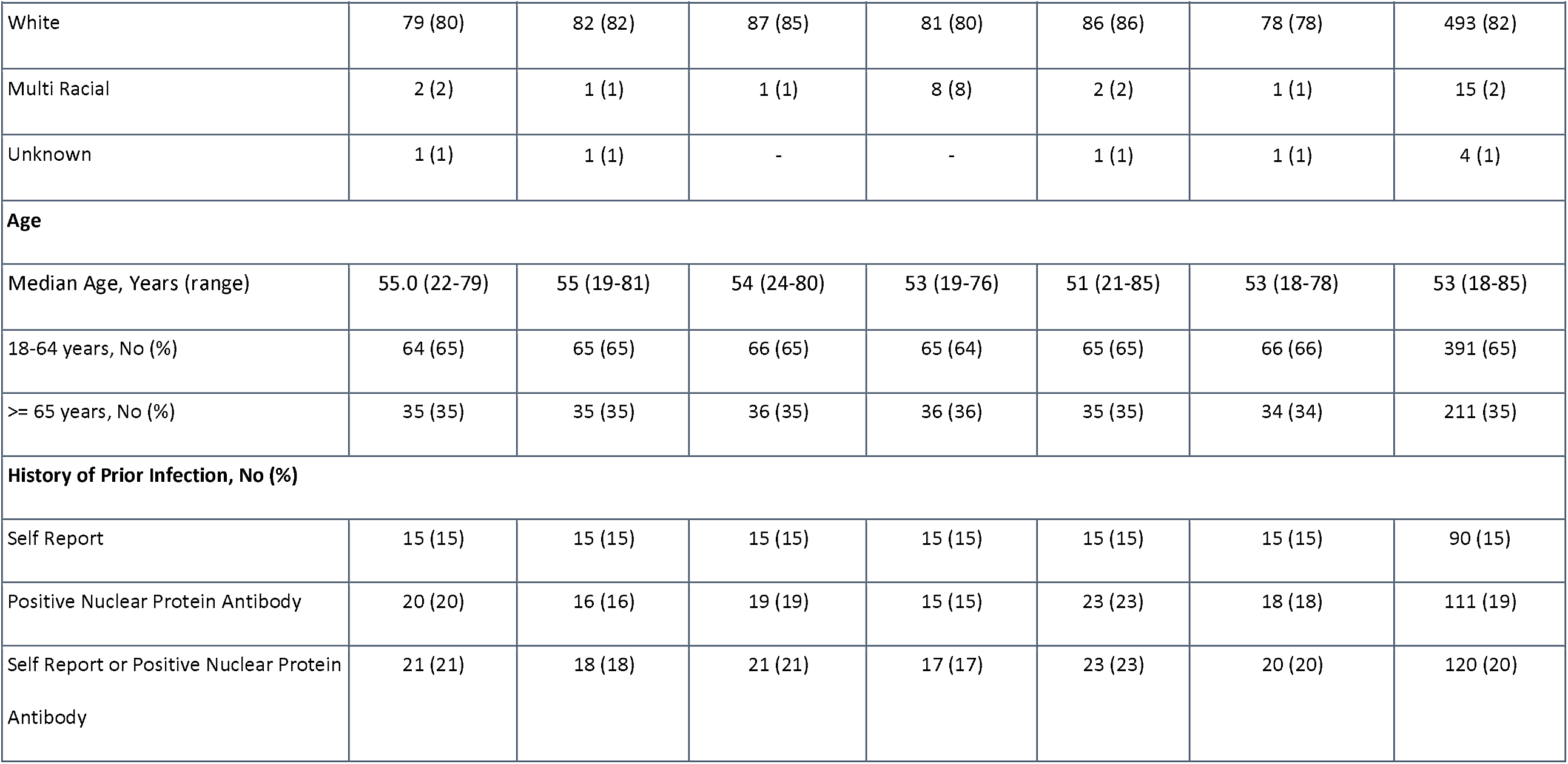
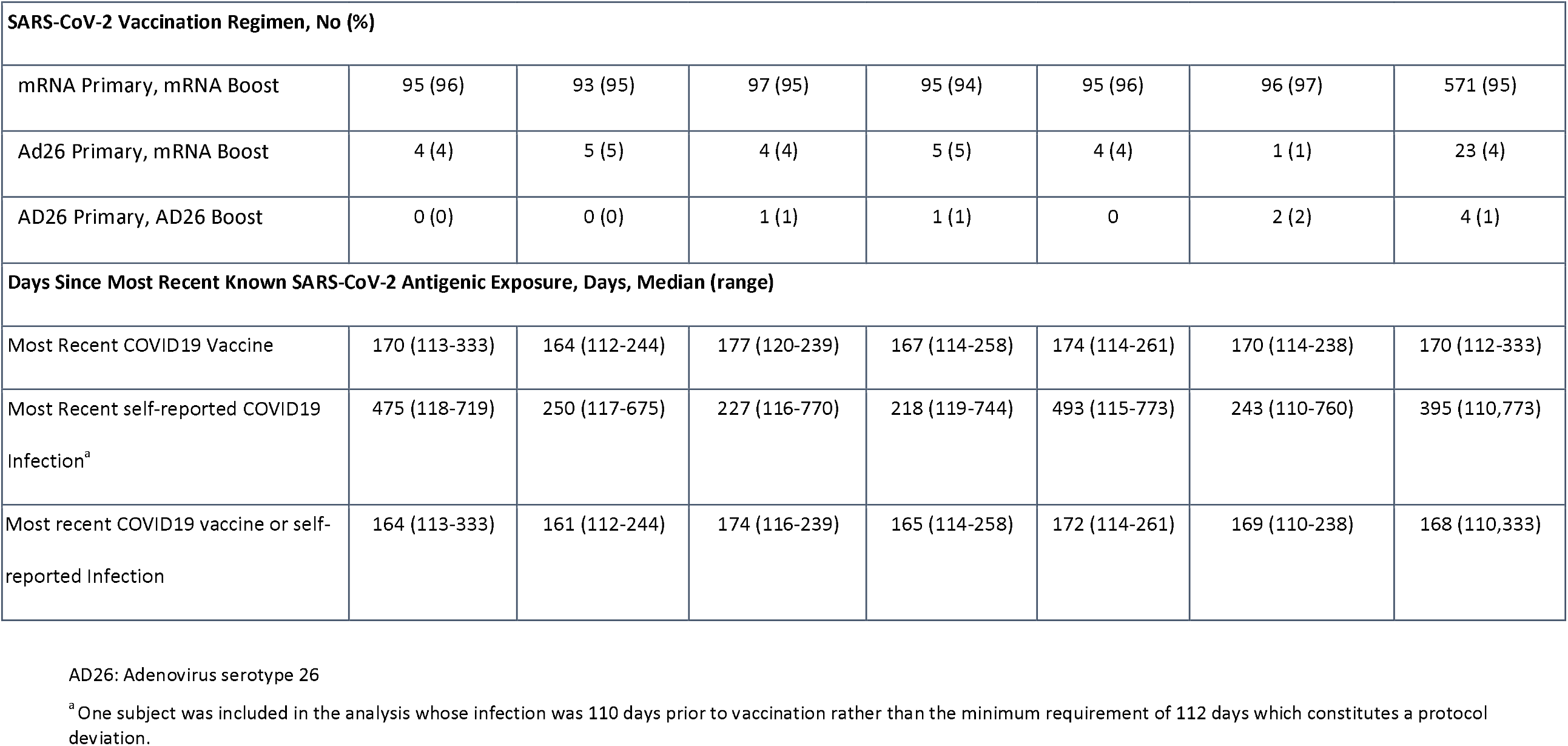
Summary of Demographic and Baseline Characteristics by Vaccination Arm – Moderna mRNA 1273 (50 mcg)

| Characteristic | ARM 1<br>1 Dose<br>Prototype<br>(N=99) | ARM 2<br>1 Dose<br>Beta + Omicron<br>BA.1<br>(N=100) | ARM 3<br>2 Dose<br>Beta+ Omicron<br>BA.1<br>(N=102) | ARM 4<br>1 Dose<br>Delta + Omicron<br>BA.1<br>(N=101) | ARM 5<br>1 Dose<br>Omicron BA.1<br>(N=100) | ARM 6<br>1 Dose<br>Omicron BA.1 +<br>Prototype<br>(N=100) | All Subjects<br>(N=602) |
| --- | --- | --- | --- | --- | --- | --- | --- |
| **Sex, No (%)** |  |  |  |  |  |  |  |
| Male Sex | 50 (51) | 43 (43) | 45 (44) | 49 (49) | 46 (46) | 52 (52) | 285 (47) |
| Female Sex | 49 (49) | 57 (57) | 57 (56) | 52 (51) | 54 (54) | 48 (48) | 317 (53) |
| **Ethnicity, No (%)** |  |  |  |  |  |  |  |
| Not Hispanic or LatinX | 94 (95) | 90 (90) | 100 (98) | 94 (93) | 93 (93) | 94 (94) | 565 (94) |
| Hispanic or LatinX | 5 (5) | 10 (10) | 2 (2) | 7 (7) | 7 (7) | 6 (6) | 37 (6) |
| **Race, No (%)** |  |  |  |  |  |  |  |
| American Indian or Alaska Native | - | 2 (2) | - | - | - | - | 2 (0) |
| Asian | 5 (5) | 6 (6) | 8 (8) | 6 (6) | 7 (7) | 9 (9) | 41 (7) |
| Native Hawaiian or Pacific Islander | - | - | - | - | - | - | - |
| Black or African American | 12 (12) | 8 (8) | 6 (6) | 6 (6) | 4 (4) | 11 (11) | 47 (8) |

| **Characteristic** | **ARM 1**<br>**1 Dose**<br>**Prototype**<br>**(N=99)** | **ARM 2**<br>**1 Dose**<br>**Beta + Omicron**<br>**BA.1**<br>**(N=100)** | **ARM 3**<br>**2 Dose**<br>**Beta+ Omicron**<br>**BA.1**<br>**(N=102)** | **ARM 4**<br>**1 Dose**<br>**Delta + Omicron**<br>**BA.1**<br>**(N=101)** | **ARM 5**<br>**1 Dose**<br>**Omicron BA.1**<br>**(N=100)** | **ARM 6**<br>**1 Dose**<br>**Omicron BA.1 +**<br>**Prototype**<br>**(N=100)** | **All Subjects**<br>**(N=602)** |
| --- | --- | --- | --- | --- | --- | --- | --- |
| White | 79 (80) | 82 (82) | 87 (85) | 81 (80) | 86 (86) | 78 (78) | 493 (82) |
| Multi Racial | 2 (2) | 1 (1) | 1 (1) | 8 (8) | 2 (2) | 1 (1) | 15 (2) |
| Unknown | 1 (1) | 1 (1) | - | - | 1 (1) | 1 (1) | 4 (1) |
| **Age** |  |  |  |  |  |  |  |
| Median Age, Years (range) | 55.0 (22-79) | 55 (19-81) | 54 (24-80) | 53 (19-76) | 51 (21-85) | 53 (18-78) | 53 (18-85) |
| 18-64 years, No (%) | 64 (65) | 65 (65) | 66 (65) | 65 (64) | 65 (65) | 66 (66) | 391 (65) |
| >= 65 years, No (%) | 35 (35) | 35 (35) | 36 (35) | 36 (36) | 35 (35) | 34 (34) | 211 (35) |
| **History of Prior Infection, No (%)** |  |  |  |  |  |  |  |
| Self Report | 15 (15) | 15 (15) | 15 (15) | 15 (15) | 15 (15) | 15 (15) | 90 (15) |
| Positive Nuclear Protein Antibody | 20 (20) | 16 (16) | 19 (19) | 15 (15) | 23 (23) | 18 (18) | 111 (19) |
| Self Report or Positive Nuclear Protein Antibody | 21 (21) | 18 (18) | 21 (21) | 17 (17) | 23 (23) | 20 (20) | 120 (20) |
| **SARS-CoV-2 Vaccination Regimen, No (%)** |  |  |  |  |  |  |  |
| mRNA Primary, mRNA Boost | 95 (96) | 93 (95) | 97 (95) | 95 (94) | 95 (96) | 96 (97) | 571 (95) |
| Ad26 Primary, mRNA Boost | 4 (4) | 5 (5) | 4 (4) | 5 (5) | 4 (4) | 1 (1) | 23 (4) |
| AD26 Primary, AD26 Boost | 0 (0) | 0 (0) | 1 (1) | 1 (1) | 0 | 2 (2) | 4 (1) |
| **Days Since Most Recent Known SARS-CoV-2 Antigenic Exposure, Days, Median (range)** |  |  |  |  |  |  |  |
| Most Recent COVID19 Vaccine | 170 (113-333) | 164 (112-244) | 177 (120-239) | 167 (114-258) | 174 (114-261) | 170 (114-238) | 170 (112-333) |
| Most Recent self-reported COVID19 Infection<sup>a</sup> | 475 (118-719) | 250 (117-675) | 227 (116-770) | 218 (119-744) | 493 (115-773) | 243 (110-760) | 395 (110,773) |
| Most recent COVID19 vaccine or self-reported Infection | 164 (113-333) | 161 (112-244) | 174 (116-239) | 165 (114-258) | 172 (114-261) | 169 (110-238) | 168 (110,333) |

AD26: Adenovirus serotype 26
<sup>a</sup> One subject was included in the analysis whose infection was 110 days prior to vaccination rather than the minimum requirement of 112 days which constitutes a protocol deviation.

### Vaccine Safety and Reactogenicity

Solicited local and systemic AEs after vaccination were comparable to other booster trials^18^ and did not differ between arms (Supplemental Figure 2). The most frequently reported solicited local AE was injection-site pain (83%). The most common solicited systemic AEs were fatigue (67%) and myalgia (58%). Most solicited AEs were mild (47%) to moderate (43%) with only 4% severe. Unsolicited AEs were similar between arms (22 to 45 events per arm). As of the data cut off, there were 4 SAEs in 3 arms, deemed unrelated to study product: dehydration, chronic obstructive pulmonary disease exacerbation, cellulitis, and a death (14 days after vaccination) from myocardial infarction in a male participant in his 70s with prior coronary artery disease. There were 18 reports of MAAEs; 2 MAAES were deemed related to study product, that have resolved or are resolving and 1 unrelated NOCMC. There were 4 AESIs reported, one of which was related to study product and has resolved. That participant, a male in his 30s, reported chest pain 1 day after vaccination that was evaluated as possible myocarditis and ultimately ruled out with a negative Troponin I and normal cardiac MRI. Symptoms subsequently resolved in 4 days. There were no AEs leading to withdrawal from the study.

### Breakthrough SARS-CoV-2 infections

There were 65 COVID-19 illnesses occurring after randomization reported by data cut off, none resulting in hospitalizations. There were 12 cases in the Prototype arm, 5 and 15 cases in the 1 and 2 dose Beta + Omicron BA.1 arms, respectively, 16 cases in the Delta + Omicron BA.1 arm, 6 cases in the Omicron BA.1 monovalent arm and 11 in the Omicron BA.1 + Prototype arm. Eleven participants with COVID-19 occurring between vaccination and Day 15 were excluded from the Day 15 immunogenicity analysis.

### Neutralizing Antibody Response

Proportions of participants with detectable neutralization antibodies for variant-containing and prototype vaccines against all tested variant viruses were 97% or greater. Baseline and Day 15 geometric mean titers (GMT) were similar between older (≥ 65 years) and younger adults in each arm, but 2-3 times higher in previously infected participants compared to uninfected participants (Supplemental Tables 4 and 5).

For uninfected participants, the Day 15 GMT against the D614G strain (GMT_D614G_) were similar across all arms (Prototype 16,976 [95% Confidence Interval (CI): 13,851-20,806]); Omicron BA.1 + Beta 16,115 [95% CI: 13,460-19,292]; Omicron BA.1 + Delta 21,972 [95% CI: 16,761-28,802]; Omicron BA.1 16,467 [95% CI: 13,087-20,719]; and Omicron BA.1 + Prototype 21,096 [95% CI: 17,398-25,580]; (Figure 1).

**FIGURE 1:**
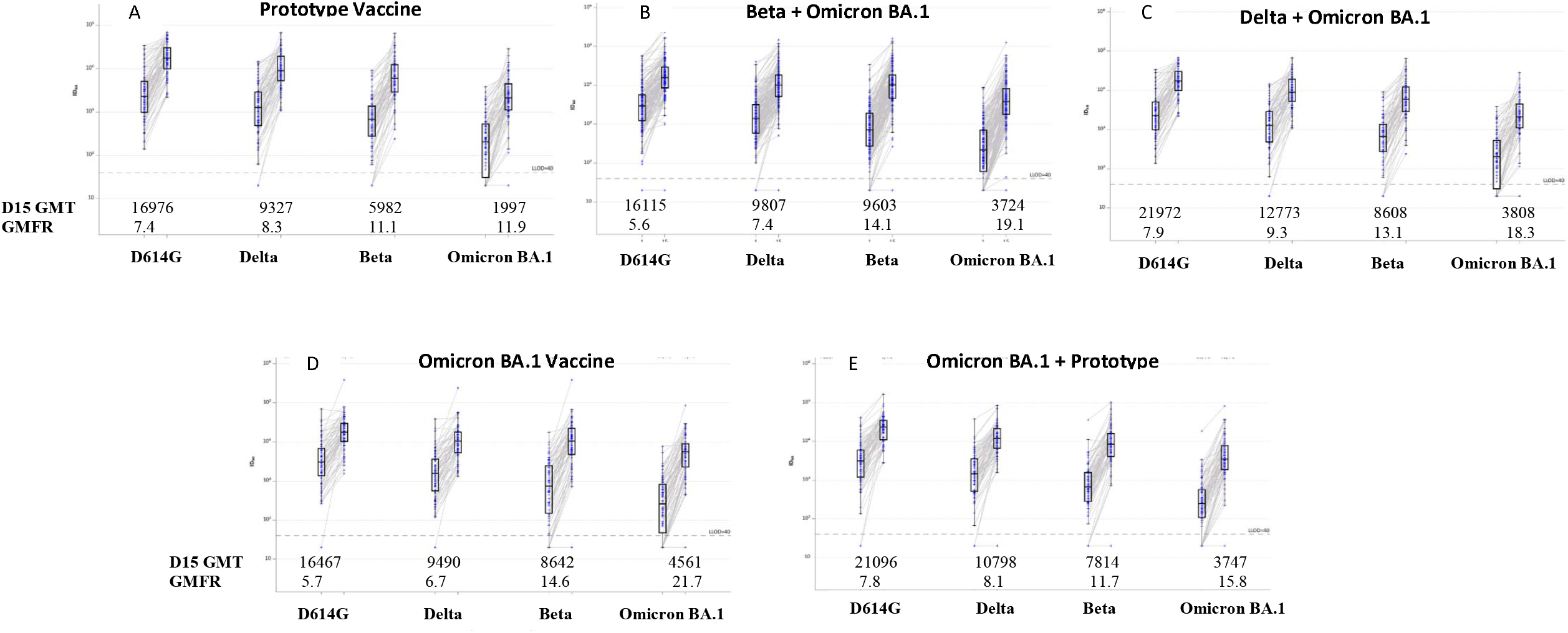
Pseudovirus Neutralization ID_50_ Titers by Timepoint (baseline and Day 15) and Variant (D614G, Delta (B.1.617.2), Beta (B.1.351) and Omicron BA.1 (B.1.1.529) in Uninfected Participants: A) Prototype monovalent vaccine, B) Omicron BA.1 + Beta bivalent vaccine C) Omicron BA.1 + Delta bivalent vaccine, D) Omicron BA.1 monovalent vaccine and E) Omicron BA.1 + Prototype bivalent vaccine. Boxes with horizontal bars denote interquartile range (IQR) and median ID_50_, respectively. Whisker denotes 95% confidence interval. LLOD, lower limit of detection of the assay. GMT, geometric mean titer at Day 15. GMFR, geometric mean fold rise from baseline to Day 15.

For uninfected participants, Day 15 GMT_BA.1_ were significantly higher following Omicron-containing vaccines than with Prototype (1,997 [95% CI: 1,482-2,692]), and similar among the different Omicron-containing vaccine arms (Omicron BA.1 + Beta 3,724 [95% CI: 3,009-4,608], Omicron BA.1 + Delta 3,808 [95% CI: 2,716-5,337], Omicron BA.1 4,561 [95% CI: 3,379-6,157], and Omicron BA.1 + Prototype 3,747 [95% CI: (2,773-5,061]). The geometric mean fold rise against Omicron BA.1 (GMFR_BA.1_) from baseline to Day 15 was 11.9 for the Prototype arm and 19.1, 18.3, 21.7 and 15.8 for the Omicron BA.1 + Beta, Omicron BA.1 + Delta, Omicron BA.1 and Omicron BA.1 + Prototype arms, respectively. Data for Delta and Beta specific GMTs and GMFRs are shown in Figure 1 and Supplemental Table 4.

In the ANCOVA model, geometric mean ratio (GMR) against the ancestral D614G strain did not differ for any of the four variant-containing vaccine candidates compared to Prototype. All Omicron-containing vaccines led to a GMR_BA.1_ of greater than 1.5 relative to the Prototyped vaccine, with unadjusted lower bound confidence interval >1; (1.71 [97.5% CI: 1.23-2.39]) for Beta + Omicron BA.1; 1.7 [97.5% CI: 1.16-2.48] for Delta + Omicron BA.1; 2.03 [97.5% CI: 1.37-3.00] for Omicron BA.1 monovalent; and 1.56 [97.5% CI: 1.06,-2.31] for Omicron BA.1 + Prototype) (Table 2).

**TABLE 2:**
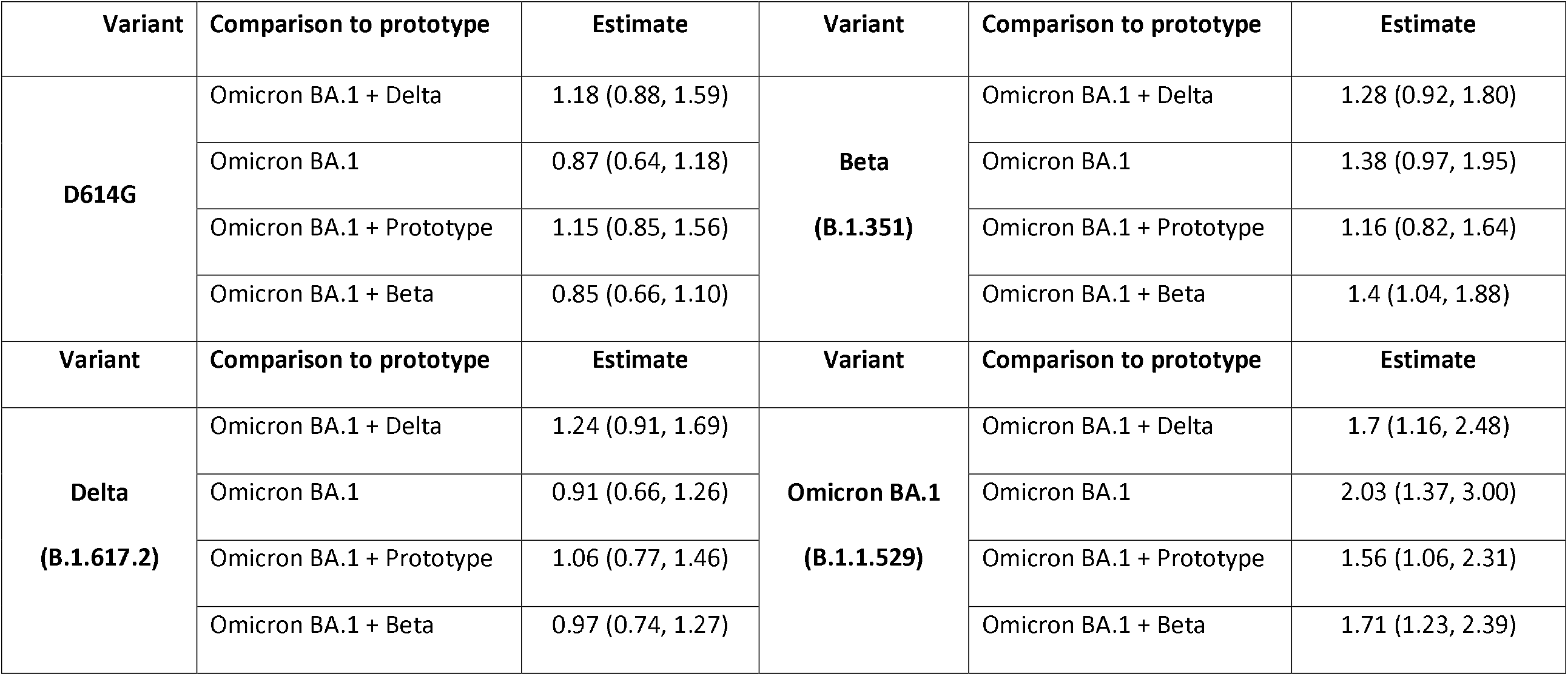
Adjusted Pseudovirus Neutralization Geometric Mean Titer at Day 15 by ANCOVA Modeling in Uninfected Participants: ANCOVA model adjusted for baseline titers and age. Confidence intervals are unadjusted at confidence level 97.5%.

A subset of 20-24 samples from uninfected participants in four arms were also tested in a different laboratory at Day 15 for neutralizing antibody titers to D614G and Omicron subvariants BA.1, BA.2.12.2 and BA.4/BA.5 (Figure 2 and Supplemental Table 5). The GMT_BA.4/BA.5_ were approximately one-third of BA.1 responses and were similar across arms (Prototype 517 [95% CI: 324-826]; Beta + Omicron BA.1 628 [95% CI:367-1,074]; Delta + Omicron BA.1 765 [95% CI: 443-1,322] and Omicron BA.1 + Prototype 635 [95% CI: 447 – 903]).

**FIGURE 2:**
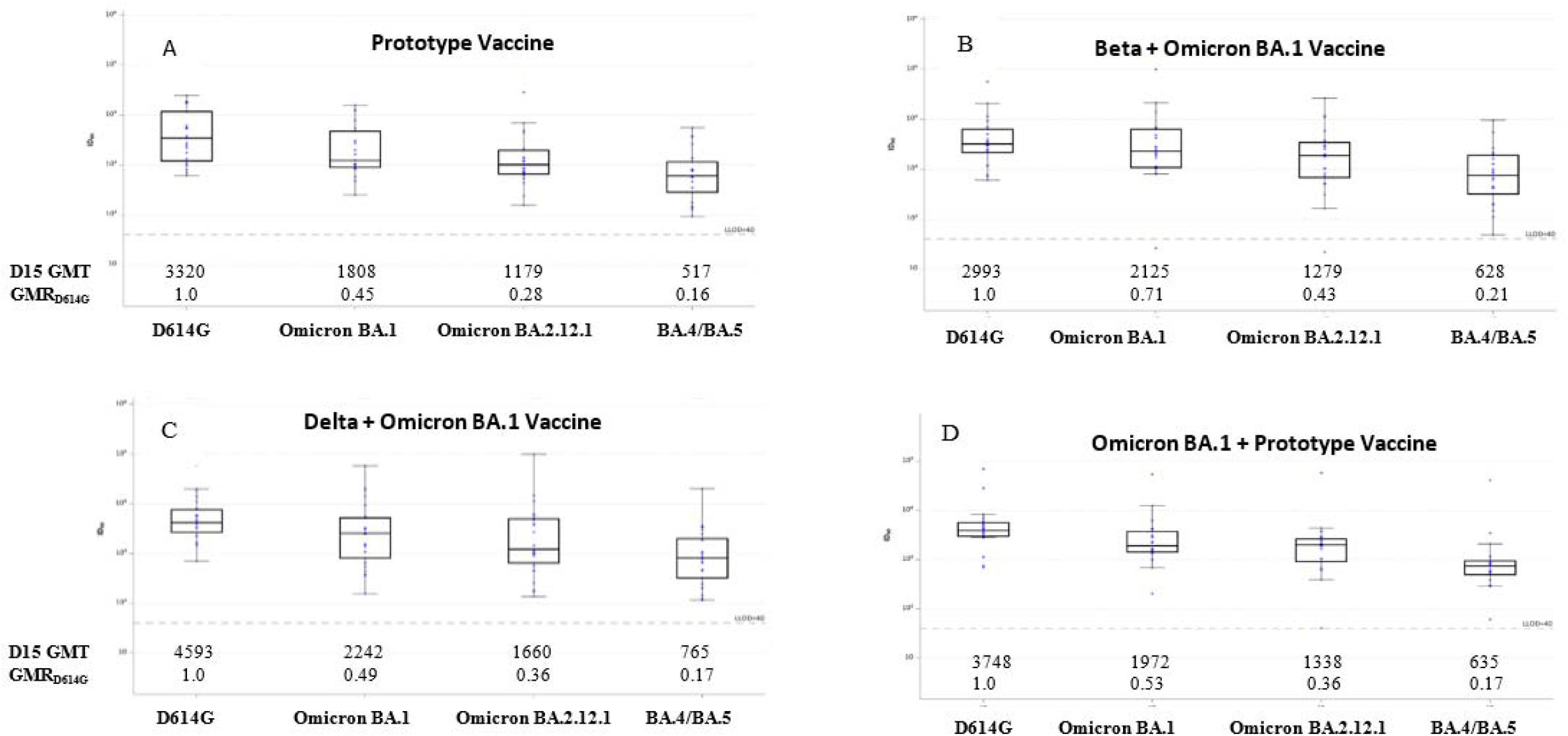
Pseudovirus Neutralization ID_50_ Titers by Timepoint (Day 15) and Variant (D614G, Omicron BA.1, Omicron BA.2.12.1, and Omicron BA.4/BA.5 in a Sample of Uninfected Participants: A) Prototype monovalent vaccine, B) Omicron BA.1 + Beta bivalent vaccine C) Omicron BA.1 + Delta bivalent vaccine, and D) Omicron BA.1 + Prototype bivalent vaccine. Boxes and horizontal bars denote interquartile range (IQR) and median ID_50_, respectively. Whisker denotes 95% confidence interval. LLOD, lower limit of detection of the assay. GMT, geometric mean titer. GMR_D614G_, geometric mean ratio against D614G.

### Antigenic Cartography and Landscapes

Figure 3A shows antigenic landscapes fitted to the GMTs of uninfected participants with the corresponding neutralizing antibody titers on the adapted base antigenic map (Figure 3B). All vaccine arms in uninfected participants had similar pre-vaccination antigenic landscapes, with the apex over D614G, as expected. After vaccination, all arms had antibody titers which raised and flattened the antigenic landscape. A second booster dose raised antibody titers in uninfected participants to the titers observed in previously-infected participants at baseline (Figure 3D-E). In both cohorts, Omicron-containing vaccines lifted titers against BA.1 the most. In the subset of vaccine candidates and samples tested against BA.4/BA.5, the Omicron BA.1 + Prototype and Omicron BA.1 + Delta vaccines generated the highest antibody titers, but Omicron BA.1 + Beta vaccinations resulted in the flattest landscapes (Figure 3C).

**FIGURE 3:**
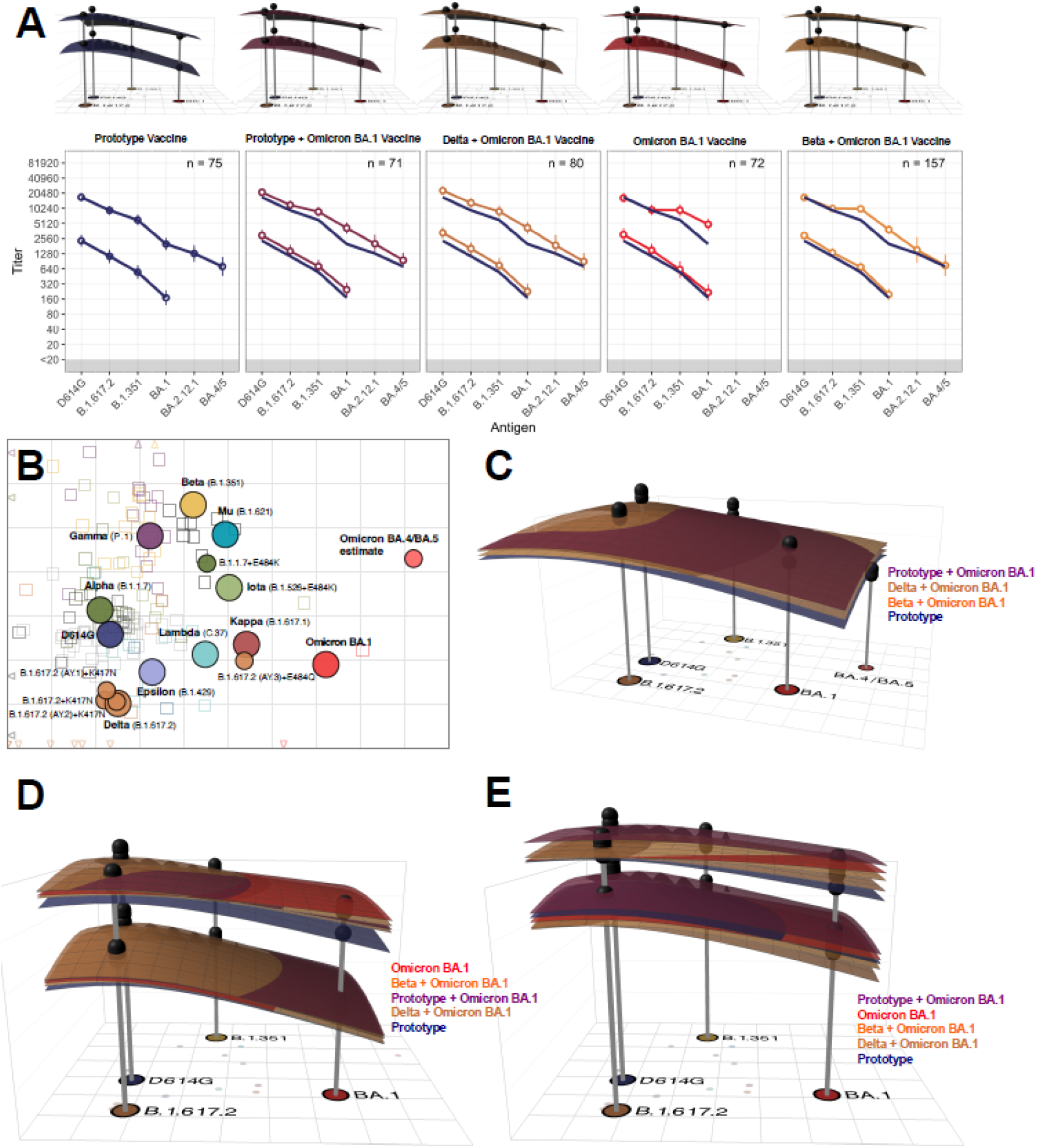
Antigenic Cartography. **A)** Day 1 and Day 15 geometric mean titer (GMT) antibody landscapes and titers for uninfected individuals in the different arms. Impulses show the GMT against the specific variant. The lower landscape and line correspond to Day 1 immunity, upper landscape to Day 15 immunity. To interpret landscapes, a Day 15 response where upper landscape is flat, indicates the responses to all the variants were equivalent, whereas, skewing up or down indicates an uneven response across variants. The titer plots show GMTs with 95% CI, where the superimposed blue line shows the Prototype GMT in each panel. **B)** An adapted version of the antigenic map by Wilks, et al. served as base map for all antibody landscapes. Virus variants are shown as filled circles, variants with additional substitutions from their root variant as smaller circles. Individual sera are displayed as open squares in the color of their root variant or grey for mRNA-1273 vaccinated sera, small dark squares represent clinical trial participants. One grid unit in the map corresponds to a two-fold dilution in the neutralization assay, within x- and y-axis the map orientation is free as antigenic distances are relative. Small triangles point to sera outside the shown map area. **C)** Day 15 GMT antibody landscapes for the arms and samples (n=87) that were titrated against BA.2.12.1 and BA.4/BA.5. The BA.4/BA.5 mean estimate shown in 3B was taken to position the titer impulse. **D)** Day 1 and day 15 GMT antibody landscapes for uninfected participants. **E)** Day 1 and day 15 GMT antibody landscapes for previously infected participants. The surface colors represent study arms (blue: Prototype, red: Omicron BA.1, purple: Prototype + Omicron BA.1, brown: Delta + Omicron BA.1, orange: Beta + Omicron BA.1). Non-responding individuals with titers <40 against all variants were excluded (n=7 uninfected, n=1 infected).

## DISCUSSION

The continued emergence of SARS-CoV-2 VOCs has led to a recommendation from both the WHO and FDA to update COVID-19 vaccines.^25,26^ Ideally, the strains selected for modified vaccines would cover currently circulating strains, including variants that drift from Omicron, like BA.4/BA.5, as well as strains that may evolve from other distinct locations on the phylogenetic tree such as occurred when Omicron BA.1 emerged and replaced the Delta variant. Therefore, it is important to investigate not only antibody titers to known variants, but also the antigenic relationships among different SARS-CoV-2 VOCs^22^ and how variant vaccines may alter immunologic landscapes to cover new strains that may emerge. Here, we described the magnitude, breadth, and landscapes of the early neutralizing antibody response following a second booster with investigational variant-specific vaccines reflective of the diverse SARS-CoV-2 immunologic background seen in the general population.

All tested vaccines were well-tolerated and induced robust serologic responses to the D614G strain as well as to Beta, Delta and Omicron BA.1 VOCs. In all arms, titers were higher against D614G compared to the VOCs, similar across age groups, and highest among those with prior infection. Day 15 ID_50_ neutralization titers against D614G for all arms exceeded titers correlated with early protection observed in the Moderna mRNA-1273 efficacy study.^27^

Although additional doses of mRNA vaccines improved neutralizing activity against Omicron BA.1, these titers remained 3-8 times lower compared to neutralization of the D614G virus; though the correlates of protection for COVID-19 caused by Omicron BA.1 are not established.^28,29^ Higher serological response against the Omicron BA.1 strain was observed with Omicron-containing vaccines compared to Prototype. Similarly, Chalkias et al. have recently shown^30^ that an mRNA bivalent vaccine (Prototype + Omicron BA.1) administered as a second boost induced significantly higher antibody responses against Omicron BA.1 compared to Prototype vaccine. In another study, a single boost of mRNA-1273 increased antibody titers against Omicron BA.1 but titers rapidly waned within 90 days after vaccination.^20^ At this point it remains unknown if an additional boost (4^th^ dose) with an Omicron-containing vaccine can alter memory specific B cell responses and slow antibody decay.^31^ Additionally, some data suggests that natural infection with earlier variants prior to the emergence of Omicron subvariants BA.4/BA.5, irrespective of vaccination history, results in marginal neutralizing antibody activity against these subvariants.^32^ This suggests that virologic escape continues to evolve and that new strategies may be needed to extend the breadth of cross-neutralizing responses, as recently recommended by the Food and Drug Administration (FDA).^26,32,33^ In our study, all vaccine candidates generated roughly one-third lower titers to BA.4/BA.5 subvariants when compared to BA.1 although detectable titers were present in 98.8% of participants tested. Due to the small number of samples tested against the Omicron BA.4/BA.5 strains, we cannot conclude that Omicron BA.1-containing vaccines will offer a serologic advantage specifically for these subvariants, although the neutralizing antibody GMT_BA.4/BA.5_ at Day 15 induced by Omicron BA.1-containing vaccines are higher than titers generated by the Prototype vaccine.

The antibody landscapes further support inclusion of variants in booster vaccines. Early after vaccination, the antigenic landscape rises and the Omicron BA.1 monovalent, Omicron BA.1 + Prototype, and Omicron BA.1 + Beta vaccines flatten the antigenic landscape more than Prototype, suggesting there may be higher titers of neutralizing antibodies with Omicron-containing variant vaccines against future VOCs if they emerge near Omicron BA.1. Notably, all landscapes dip the furthest over Omicron BA.4/BA.5 suggesting the least protection in that antigenic space.

Our study has some limitations. First, the sample size is small for participants with prior infection (20%) and adults older than 65 years (35%). Stratification by prior vaccine regimen or interval since last antigenic exposure was limited by the sample size. Second, we are presenting interim virus neutralization titer results only through Day 15, and antibody titers may not peak until Day 29, though this is unlikely to change the interpretation. Planned later timepoints will provide important data on durability of the response. Third, T cell responses, which may be critical to preventing severe disease, have not yet been evaluated. Finally, only Moderna vaccines were studied in this initial stage. Later stages are assessing Pfizer/BioNTech mRNA and GSK/Sanofi protein-adjuvanted variant vaccine candidates.

Recently, the FDA recommended that manufacturers include Omicron BA.4/BA.5 along with Prototype in fall 2022 SARS-CoV-2 vaccine formulations.^26^ While at this time there are no clinical data with a BA.4/BA.5 vaccine to indicate how this variant vaccine may alter antigenic landscapes. Our data are consistent with the FDA’s recommendation with a vaccine with a prototype component to increase antigenic landscapes near D614G in case new VOCs emerge in that part of the antigenic map, and an Omicron component to increase antigenic landscapes near Omicron BA.1 and BA.4/BA.5. It is recognized that all strategies tested and reported here provide suboptimal coverage of the antigenic space near Omicron BA.4/BA.5. If future VOCs are similar to BA.4/BA.5 but continue to increase antigenic distance from D614G, optimizing the antigenic landscape near BA.4/BA.5 will be critical to improve COVID-19 vaccine effectiveness.

## Data Availability

All data produced in the present study are available upon reasonable request to the authors

## ACKNOWLEDGEMENTS

We thank all the participants in this trial; the members of the safety monitoring committee (Kawsar Talaat, MD, John Treanor, MD, Grant Paulsen, MD and Donald Stablein, PhD), who provided thoughtful discussions resulting in the early trial design; and Moderna for their collaboration, scientific input, and sharing of documents needed to implement the trial.

The COVAIL trial has been funded in part with federal funds from the National Institute of Allergy and Infectious Diseases (NIAID) and the National Cancer Institute (NCI), National Institutes of Health (NIH), under contract HHSN261200800001E 75N910D00024, task order number 75N91022F00007. This work was also supported in part with federal funds from the NIAID, NIH under contract number 75N93021C00012 and by the Infectious Diseases Clinical Research Consortium (IDCRC) through NIAID under award number UM1AI148684.

DJ Smith, A Netzl, SH Wilks and S Türeli were supported by the NIH NIAID Centers of Excellence for Influenza Research and Response (CEIRR) contract 75N93021C00014 as part of the SAVE program. D Montefiori and A Eaton were supported by the NIAID Collaborative Influenza Vaccine Innovation Centers (CIVICs) contract 75N93019C00050.

Testing of neutralizing antibody titers by Monogram Biosciences, Labcorp has been funded in part with federal funds from the Department of Health and Human Services; Office of the Assistant Secretary for Preparedness and Response; Biomedical Advanced Research and Development Authority, under Contract No. 75A50122C00008. Testing for anti-N specific antibody was conducted by Cerba Research under Contract No. 75N93021D00021.

The manuscript content is solely the responsibility of the authors and does not necessarily represent the official views of the National Institute of Allergy and Infectious Diseases, the National Institutes of Health.

## DISCLOSURES

ARB has research grants from Pfizer, Janssen, Merck Sharp and Dohme and CyanVac and serves on the DSMB board for AzurX Biopharma and has served as a consultant for Janssen. ARF has research grants from Pfizer, Janssen, CyanVac, Merck Sharp and Dohme and BioFire Diagnostics and serves on the DSMB for Novavax. EW has research grants from Janssen, Merck Sharp and Dohme and Pfizer and served on a DSMB sponsored by GlaxoSmithKline. NGR has research grants from Pfizer, Sanofi Pasteur, Merck Sharp and serves on the Safety Monitoring Committees for ICON and EMMES. DSG receives consulting fees from Critica, Inc a non-profit organization. LRB has received grant support from NIH for SARS-CoV-2 therapeutic and vaccine development work and is involved in SARS-CoV-2 vaccine trials conducted in collaboration with the NIH, Covid Prevention Network (CoVPN), Crucell/Janssen, Moderna, and Harvard Medical School. SRW has conducted clinical trials funded by Sanofi Pasteur, Janssen Vaccines, ModernaTx, and NIAID/NIH and chairs an Independent Data Monitoring Committee (IDMC) for Janssen Vaccines. CMH is a member of a Safety Monitoring Committee for the Emmes Corporation. SJL has received research grant support to UCSD from Gilead Sciences. Her institution has also received funding from NIH to conduct clinical trials of Janssen and AstraZeneca COVID-19 vaccines. EJA has consulted for Pfizer, Sanofi Pasteur, Janssen, GSK, Moderna, and Medscape, and his institution receives funds to conduct clinical research unrelated to this manuscript from MedImmune, Regeneron, PaxVax, Pfizer, GSK, Merck, Sanofi-Pasteur, Janssen, and Micron. He also serves on data and safety monitoring boards for Kentucky BioProcessing, Inc. and Sanofi Pasteur. He also serves on safety monitoring committees for WCG-ACI Clinical. His institution has also received funding from NIH to conduct clinical trials of Moderna and Janssen COVID-19 vaccines. SK’s institution has received funding from CDC to conduct clinical research unrelated to this manuscript, from NIH to conduct clinical trials of Moderna and Janssen COVID-19 vaccines, and from Pfizer to conduct clinical trials of Pfizer COVID-19 vaccine. CAR’s institution has received funds to conduct clinical research unrelated to this manuscript from BioFire Inc, GSK, MedImmune, Micron, Janssen, Merck, Novavax, PaxVax, Regeneron, Sanofi-Pasteur. She is co-inventor of patented RSV vaccine technology unrelated to this manuscript, which has been licensed to Meissa Vaccines, Inc. Her institution has received funds from NIH, Moderna, Pfizer, and Janssen to conduct clinical trials of COVID-19 vaccines. EBW has served as an investigator for vaccine studies sponsored by Pfizer, Moderna and Sequiris. He has served on advisory boards for Vaxcyte and Iliad Biotechnologies. MJS has served as an investigator for vaccine studies sponsored by Pfizer, and has received investigator-initiated grants from Merck related to antimicrobial stewardship. RMN has received research grant funding from ModernaTX, Inc and Janssen Vaccines & Prevention B.V. AW has received funding from NIH, GlaxoSmithKline and Sanofi and has been a consultant for Aicuris, X-Vax, Vir, Crozet, and Auritec. ACK reported clinical trials contract funding for vaccines or MAB vs SARS-CoV-2 with Regeneron and Pfizer. AFL has received research grant support to UCSF from Astra Zeneca. The Washington University Clinical Trials Unit has received unrelated funding support in sponsored research agreements from Moderna. PAG receives grant funding from the NIH and is a consultant for Johnson and Johnson. LCI has received research grant support from NIH, CDC, and serves on the North American Advisory Board for Moderna, Inc. LCI’s institution has received funds to conduct clinical trials of Novavax and Moderna CoVID-19 Vaccines in collaboration with NIH, US CoVID-19 Prevention Network (CoVPN). LCI’s institution has received funds to conduct clinical research unrelated to this manuscript from Merck, GSK, Novavax, and Pfizer. MJM reported laboratory research and clinical trials contract funding for vaccines or MAB vs SARS-CoV-2 with Lilly, Pfizer, and Sanofi; personal fees for Scientific Advisory Board service from Merck, Meissa Vaccines, Inc. and Pfizer.

## Notes

### Clinical Trial

NCT05289037

### Funding Statement

The trial was sponsored and funded by the National Institute of Allergy and Infectious Diseases.

### Author Declarations

The trial was reviewed and approved by the Advarra central institutional review board which gave ethical approval for this work.

