## supplemental appendix for "SARS-CoV-2 Variant Vaccine Boosters Trial: Preliminary Analyses"

This appendix is submitted by the authors to provide additional information about their work.

Supplementary Appendix to Manuscript Entitled

**SARS-CoV-2 Variant Vaccine Boosters Trial: Preliminary Analyses.**

Statistical Methods ……………………………………………………………………………………………………….………..19
Supplemental Tables
Table S1. List of COVAIL US Sites……………………………………………………………………………………………..21

Table S2. Eligible Regimens for Primary Vaccination Series and Boost…………………………………..…23
Table S3. Study Arms……………………………………………………………………………………………………………….24

Table S4. Pseudovirus Neutralization ID_50_ Titers, by Timepoint (baseline and Day 15) and Variant (D614G, Delta (B.1.617.2), Beta (B.1.351) and Omicron BA.1 (B.1.1.529) in All, Uninfected and Infected Participants in all Arms..............................................................................................…..25
Table S5. Pseudovirus Neutralization ID_50_ Titers, by Timepoint (baseline and Day 15) and Variant (D614G, Omicron (B.1.1.529, BA.2.12.1. and BA.4/BA.5) in All and Uninfected Participants in all Arms…………………………................................................................................................................ 30
Supplemental Figures
Figure S1: Consort Diagram for Stage 1......................................………………………………………………32

COVAIL Manuscript Study Group

**George Washington University, Washington D.C.**

David J. Diemert, MD; Elissa Malkin, DO; Jeffrey M. Bethony, PhD; Aimee Desrosiers, PA-C; Marc Siegel, MD

**University of Rochester VTEU, Rochester, NY**

Angela R. Branche, MD, Ann R. Falsey, MD, Edward Walsh, MD, Patrick Kingsley, BS, Michael Peasley, BS

**Emory University Hope Clinic, Decatur, GA**

Nadine G. Rouphael, MD; Cecilia Losada, MD; Daniel S. Graciaa, MD; Hady Samaha, MD; Cassie Grimsley Ackerley, MD; Kristen E. Unterberger, PA

**Brigham and Women’s Hospital, Harvard Medical School, Boston, MA**

Lindsey R. Baden, MD; Amy C. Sherman, MD; Stephen R. Walsh, MD; Alexandra Tong, BS; Rebecca Rooks, BS

**Saint Louis University, St. Louis, MO**

Sharon E. Frey, MD; Getahun Abate, MD, PhD; Zacharoula Oikonomopoulou, MD; Daniel F. Hoft, MD, PhD; Irene Graham, MD

**Departments of Molecular Virology and Microbiology and Medicine, Baylor College of Medicine, Houston, TX**

Jennifer A. Whitaker, MD; Hana M. El Sahly, MD; Wendy A. Keitel, MD; C. Mary Healy, MD

**Department of Medicine, Division of Infectious Diseases and Global Public Health, University of California San Diego, La Jolla, CA**

Susan J. Little, MD; Thomas C.S. Martin, MD; Nicole Carter, MPH; Steven Hendrickx, RN

**Center for Childhood Infections and Vaccines (CCIV) of Children’s Healthcare of Atlanta and Emory University Department of Pediatrics, Atlanta, GA**

Evan J. Anderson, MD; Christina A. Rostad, MD; Satoshi Kamidani, MD; Etza Peters, RN

**Duke Human Vaccine Institute, Duke University School of Medicine, Durham, NC**

Emmanuel B. Walter MD, MPH; Michael J. Smith MD, MSCE; M. Anthony Moody, MD; Kenneth E. Schmader, MD

**University of Illinois at Chicago-Project WISH, Chicago, IL**

Richard M. Novak, MD; Benjamin G. Ladner, MD; Andrea Wendrow, RPh; Jessica Herrick, MD

**University of Texas Medical Branch, League City, TX**

Richard Rupp, MD; Laura Porterfield, MD

**Kaiser Permanente Washington Health Research Institute, Seattle, WA**

Lisa A. Jackson, MD, MPH; Maya Dunstan, MS, RN; Rebecca Lau, PharmD; Barbara Carste, MPH

**Departments of Medicine, Epidemiology, and Laboratory Medicine & Pathology, University of Washington, Vaccines and Infectious Diseases Division, Fred Hutchinson Cancer Center, Seattle, WA**

Tara M. Babu, MD, MSCI; Anna Wald, MD, MPH; Taylor Krause, BA; Kirsten Hauge, MPH

**NYU VTEU Manhattan Research Clinic at NYU Grossman School of Medicine, New York, NY**

Angelica C. Kottkamp, MD; Tamia Davis, NP; Celia Engelson, NP; Vijaya Soma, MD

**Zuckerberg San Francisco General, University of California San Francisco, San Francisco, CA**

Anne F. Luetkemeyer, MD; Chloe Harris, BA; Azquena Munoz Lopez, BS

**Morehouse School of Medicine, Atlanta, GA**

Lilly C. Immergluck, MD; Erica Johnson, PhD; Austin Chan, MD

**Washington University School of Medicine, St. Louis, MO**

Rachel M. Presti, MD, PhD; Jane A. O’Halloran, MD, PhD; Ryley M. Thompson

**NYU VTEU Long Island Research Clinic at NYU Long Island School of Medicine, Mineola, NY**

Martín Bäcker, MD; Kimberly Byrnes, RN; Asif Noor, MD

**University of Iowa College of Medicine, Iowa City, IA**

Patricia L. Winokur, MD; Jeffery Meier, MD; Jack Stapleton, MD

**Howard University College of Medicine, Howard University Hospital, Washington D.C.**

Siham M. Mahgoub, MD; Celia Maxwell, MD; Sarah Shami, PharmD

**University of Alabama at Birmingham, Birmingham, AL**

Paul A. Goepfert, MD

**Tulane University School of Medicine, New Orleans, LA**

Dahlene N. Fusco, MD; Arnaud C. Drouin, MD; Florice K. Numbi, MD

**IDCRC Principal Investigators**

David S. Stephens, MD; Kathleen M. Neuzil, MD

**IDCRC Leadership Operations Center**

Monica M. Farley, MD; Jeanne Marrazzo, MD; Sidnee Paschal Young

**IDCRC Clinical Operations Unit**

Jeffery Lennox, MD; Robert L. Atmar, MD; Linda McNeil FHI360

**IDCRC Statistical and Data Science Unit**

Elizabeth Brown, PhD

**IDCRC Laboratory Operations Unit – Fred Hutchinson Cancer Center, Seattle, WA**

Christine M. Posavad, PhD; Megan A. Meagher, BS; Julie McElrath, MD; Mike Gale, PhD

**The Emmes Company, LLC, Rockville, MD**

Mat Makowski, PhD; Heather Hill, MS; Jim Albert, MS, Holly Baughman; Lisa McQuarrie, MS; Kalyani Telu, MS; Jinjian Mu, PhD

**Clinical Monitoring Research Program Directorate, Frederick National Laboratory for Cancer Research, Frederick, MD**

Teri C. Lewis, BS; Lisa A. Giebeig, MS; Theresa M. Engel, MFS; Caleb J. Griffith, MPH; Wendi L. McDonald, BSN; Alissa E. Burkey, MS; Lisa B. Hoopengardner, MS; Jessica E. Linton, MS; Nikki L. Gettinger, MPH

**Division of Microbiology and Infectious Diseases, National Institute of Allergy and Infectious Diseases, National Institutes of Health, Bethesda, MD.**

Marina Lee, PhD; Mamodikoe Makhene, MD; Mohamed Elsafy, MD; Rhonda Pikaart-Tautges, BS; Janice Arega, MS; Binh Hoang, RPh; Dan Curtin; Hyung Koo, BSN; Elisa Sindall, BSN; Sonja Crandon, BSN; Seema U. Nayak, MD; Marciela M DeGrace, PhD; Diane J Post, PhD; Paul C Roberts, PhD; John H Beigel, MD.

COVAIL Manuscript Study Team Members

**George Washington University, Washington D.C.**

David J. Diemert, MD; Elissa Malkin, DO; Jeffrey M. Bethony, PhD; Aimee Desrosiers, PA-C; Marc Siegel, MD; Nikita Schroll-McLaughlin, MS; Jonathan Manning, BA; Jane Ryu, MS; Hanna-Grace Rabanes, MPH; Khadija Khan, MPH; Laura Vasquez, MPH; Caroline Thoreson, PA-C; Larissa Scholte, PhD; Rafaela Thur, DVM; Peyton St. John, BS; Dorinne Mettle-Amuah, PharmD

**University of Rochester VTEU, Rochester, NY**

Angela R. Branche, MD, Ann R. Falsey, MD, Edward Walsh, MD, Patrick Kingsley, BS, Arthur Zemanek, BSN, MS Katherine Elena, BSN, Spencer Obrecht, BSN, Ian Shannon, BSN, Amy Kaychalo, BS, MS, Erin Nowicki, Sharon Moorehead, Kari Steinmetz, BA, Doreen Francis, RN, Tanya Smith, BS, William Hamilton, BS, Jeanne Holden-Wiltse, MPH, MBA, Christopher Lane, MS, Michael Peasley, BS, Samuel Diehl, BS, Kyle Richards, PharmD, Stephen Bean, PharmD, Nicole Dornbush, PharmD, Carol Cole, PharmD

**Emory University Hope Clinic, Decatur, GA**

Nadine G. Rouphael, MD; Cecilia Losada, MD; Daniel S. Graciaa, MD; Hady Samaha, MD; Cassie Grimsley Ackerley, MD; Kristen E. Unterberger, PA; Amy Anderson, BSN; Mary Atha, ACNP; Kareem Bechnak, BSN; Sarah Bechnak, BSN; Mary Bower, BSN; Laura Clegg, RN; Matthew Collins, MD, PhD; Francine Dyer, RN; Srilatha Edupuganti, MD; Rebecca Fineman, BS; Tigisty Girmay, MSN; Rebecca Gonzalez, PharmD; Natalie Gray, BS; Evan Gutter, MPH; Lisa Harewood; Chris Huerta, MSc; Brandi Johnson, BS; Lauren Johnson, MPH; Colleen Kelley, MD; Alexandra Koumanelis, BA; Deborah Laryea, BSN; Hollie Macenczak, BSN; Nour Makkaoui, MD; Michele McCullough, MPH; Tuong-Vy Ngo, PharmD; Eileen Osinski, BS; Julia Paine, BS; Bernadine Panganiban, BS; Rose Pope, RN; Paulina Rebolledo, MD; Susan Rogers, RPh; Erin Scherer, PhD; Veronica Smith, NP-C; Andre Stringer, BS; Jessica Traenkner, PA; Dongli Wang, BS; Alahna Watson, BA; Stacey Wheeler, RN; Jean Winter; Jianguo Xu, PhD

**Brigham and Women’s Hospital, Harvard Medical School, Boston, MA**

Lindsey R. Baden, MD; Amy C. Sherman, MD; Stephen R. Walsh, MD; Alexandra Tong, BS; Rebecca Rooks, BS; Jane A. Kleinjan, NP; Jon A. Gothing, NP; Andres A. Avila Paz, BA; Muneerah M. Aleissa, PharmD, MPH; Bethany Evans, BA, August Heithoff, BS; Natalie E. Izaguirre, MS; Hannah Jin, MPH; Urwah Kanwal, BS; Austin Kim, BS; Julia E. Klopfer, BS; Christina Montesano, BS; John Almeida, BA; Emily S. Koleske, BS; Hannah Levine, BS; Nicholas P. Morreale, BS; Omolola Ometoruwa, BS; Jun Bai Park Chang, BS; Anna F. Piermattei, BA; Djenane M. Pierre, BS; Megan Powell, BA; Kevin Zinchuk, PharmD; Stephanie Pickford, PharmD; Charles M. Kelly III, PharmD; Xiaofang Li, PhD; John Kupelian, BS ; Kimberly Dufresne, BS; Xiaoguang Fan, MD, PhD; Xi Zhang, PhD; Esther Arbona-Haddad, MD; Jose Humberto Licona, MD

**Saint Louis University, St. Louis, MO**

Sharon E. Frey, MD; Getahun Abate, MD, PhD; Zacharoula Oikonomopoulou, MD; Daniel F. Hoft, PhD, MD; Irene Graham, MD; Azra Blazevic, DVM, MPH; Tamara Blevins, MS; Kathleen Chirco, BSN; Sabrina M. DiPiazza, BSN, MA; Stanley Doublin; Heather Hoertel Douds, MSNS, BSN; Carol G. Duane, PhD, RN; Eric Eggemeyer, BA; Linda M. Eggemeyer-Sharpe, BSN; Lauren Nicole Foreman, BSN; Sarah Louise George, MD; Geoffrey J. Gorse, MD; Michelle Harris, PharmD; Helay Hassas, PharmD; Rong Hou, MD; Ryan Clark Kerr, BSN; Kate Elizabeth Liefer, BSN; Melissa J. Loyet, RN; Lainey Mejia-Jauregui, BS; Keith Meyer, BS; Tracy Renee Montauk, BSN; Karla J. Mosby, RN; Amanda Nethington, BS; Huan Ning, MD; Nicole Purcell; Joan M. Siegner, BSN, MA; Janice M. Tennant, BSN, MPH; Mei Xia, PhD; Kiana Wilder, BA; Cassandra Nicole Zehenny, BSN

**Departments of Molecular Virology and Microbiology and Medicine, Baylor College of Medicine, Houston, TX**

Jennifer A. Whitaker, MD; Hana M. El Sahly, MD; Wendy A. Keitel, MD; C. Mary Healy, MD; Robert L. Atmar, MD; Pedro A. Piedra, MD; Jesus Banay; Kathy Bosworth; Janet Brown, RPh; Kayla Burrell; Jeremy Castro; Tykel Eddy; Marcena Eubanks; Cathy Faw, RPh; Rachel Froebe; Alix Halter, RN; Janey John, MSN, APRN, FNP-C; Chanei Henry, AAS; Vanessa Martinez; Carol Mundell, RN; Brandie Phillips, RN; Alicia Prevost-Barthe, RN; Connie Rangel, RN; Yolanda Rayford, MS; Yvette Rugeley; Maria Shlyapobersky; Tina Sierra; Elizabeth Silguero; Lisreina Toro; Dawn Turner, RN; Chianti Wade-Bowers, RN; Jessica Woods, RN

**Department of Medicine, Division of Infectious Diseases and Global Public Health, University of California San Diego, La Jolla, CA**

Susan J. Little, MD; Thomas C.S. Martin, MD; Nicole Carter, MPH; Steven Hendrickx, RN; Ajay Bharti, MD; Alyssa Phillips; Aurora Verduzco Gonzalez, NP; Cheryl Dullano; Chris Houston; Dawn Rosenblum, RN; DeeDee Pacheco; DeLys Brooks; Fang Wan; Helene Le, CPhiT; JC Alcantar; Jill Blumenthal, MD; Joseph Lencioni, MABMH; Kory Hess; Letty Muttera, PharmD; Marlene Arredondo; Megan Smyth; Megan Taylor; Melinda Stafford, PharmD; Michelle Orsburn, MD; Michelle Truong; Niamh Higgins, PharmD, MSc, AAHIVP; Nimish Patel, PharmD, PhD, AAHIVP; Rebecca Gonzalez; Vivian Maldonado

**Center for Childhood Infections and Vaccines (CCIV) of Children’s Healthcare of Atlanta and Emory University Department of Pediatrics, Atlanta, GA**

Evan J. Anderson, MD; Christina A. Rostad, MD; Satoshi Kamidani, MD; Etza Peters, RN; Larry Anderson, MD; Julia Bartol; Leisa Bower RN; Natsuko Campbell RN; Lisa Harewood, Hui-Mien Hsiao, Laila Hussaini MPH, Inara Jooma, Gidget Kettle RN; Marcia Lewis RN; Wensheng Li; Cindy Lubbers RN; Lisa Macoy RN; Molly Morrison, Heather Nurse RN; Anna Siaw-Anim; Kathleen Stephens RN; Madeline Taylor; Ashley Tippett MPH

**Duke Human Vaccine Institute, Duke University School of Medicine, Durham, NC**

Emmanuel B. Walter MD, MPH; Michael J. Smith MD, MSCE; M. Anthony Moody, MD; Kenneth E. Schmader, MD; Susan Doyle; Lynn S Harrington BSN; Lori Hendrickson BSN; Amy O’Berry MSN; Sherry Huber BSN; Janet Wootton RN, RSCN; Kelly Clark BA; Lani Banez; Stephanie Smith BA; Byron Hauser BS; Ally Odom BA; Emily Randolph BA; Krystina Yoder BA; Kathlene Chmielewski; Luis Ballon BA; Aubree Latorre; Breana Montgomery; Antony Tritz MS; Thad Gurley, MS; Margaret Pendzich

**University of Illinois at Chicago-Project WISH, Chicago, IL**

Richard M. Novak, MD; Benjamin G. Ladner, MD; Andrea Wendrow, RPh; Jesica Herrick, MD; Alfredo J. Mena Lora, MD; Scott A. Borgetti, MD; Diana L Bahena, APRN; Regina Harden, BA; Renyce Powell;  David C. M. Chan, PharmD; Rebeca F. Gasari, PharmD; Michael Pacini, PharmD; Margarita M. Villarreal, CPhT; Rodrigo Reyes, ADN; Samuel M. Rene, MPH; Shannon M Whitted, BSN; Habiba Sultana, MBBS; Nanu Kunwar, BS; Tasmin Sultana, MBBS; Md R. Amin, PhD;  Mahmood Ghassemi, PhD,  Liam Morrissy, BS; Nia O'Neal, BS; Chasity Serrano, BS; Charlie Peterson, BA

**University of Texas Medical Branch, League City, TX**

Richard Rupp, MD; Laura Porterfield, MD; Amber Stanford, PA-C; Robert Cox, RN; Kristin Pollock, RN; Diane Barrett, MS; Gerrianne Casey, RN; Amy McMahan, LVN; Cori Burkett, PA-C; Essie Cox

**Kaiser Permanente Washington Health Research Institute, Seattle, WA**

Lisa A. Jackson, MD, MPH; Maya Dunstan, MS, RN; Rebecca Lau, PharmD; Barbara Carste, MPH; Wesley A. Andersen, RPh, MHA, MA; Lee Barr, RN; Jesse Berg, BS; Cassandra Bryant, BS; Joe Choe, BS; Jana ffitch, LPN; Lynn Gross, PA-C; Erika Kiniry, MPH; Bonnie Y Lam, PharmD; De Vona Lang; Jasmine S. Lau, BS; Stella Lee, BA; Paula J Lins, PA-C, MPH; Amy Mohelnitzky, PA-C; Marilyn Nguyen, BS; Matthew Nguyen, MPH; Hallie Phillips, MEd; Stephanie Pimienta, BS; Melissa Resendiz Rivas, BA; Melissa Boothe Scheer, PA-C; Janice Suyehira, MD; Stacie Wellwood, LPN; Maryann K Woodford, PA-C

**Departments of Medicine, Epidemiology, and Laboratory Medicine & Pathology, University of Washington, Vaccines and Infectious Diseases Division, Fred Hutchinson Cancer Center, Seattle, WA**

Tara M. Babu, MD, MSCI; Anna Wald, MD, MPH; Taylor Krause, BA; Kirsten Hauge, MPH; Jina Taub, ARNP, Dana Varon, ARNP, Britt Murphy, ARNP, Morissa Pertik, PA-C, T. Nui Pholsena, ARNP, Alyssa Braun, BS, Mark Drummond, BS, Jessica Heimonen, MPH, Amy Link, BS, Lindsey McClellan, BS, Jessica Moreno, BS, Chloe Wilkens, BS, Matt Seymour, MPH, Lawrence Hemingway, BS, Jean Mernaugh, BS, Chris McClurkan, BS, Kerry Laing, PhD, Meredith Potochnic, PharmD, Joong Kim, PharmD, Bao-Chao Vo, PhT, Dil Singh, BS

**NYU VTEU Manhattan Research Clinic at NYU Grossman School of Medicine, New York, NY**

Angelica C. Kottkamp, MD; Tamia Davis, NP; Celia Engelson, NP; Vijaya Soma, MD; Abdulwahab Abdulai; Ashanay Allen; Natella Aronova, NP; Philip Aziz, PharmD; Emily Beato; Samuel Bliss, PharmD; Jacqueline Callahan, RN; Ellie Carmody, MD; Aimee Edwin, RN; Shelby Goins; Sarah Haiken; Ramin Herati, MD; Abdonnie Holder; Janice Hong; Trishala Karmacharya; Manpreet Kaur, PharmD; Hye-Youn Kim; Alexander McMeeking, MD; Mark Mulligan, MD; Wai Ng; Edward Nirenberg; Samuel Nweke; Lalitha Parameswaran, MD; Levonne Phillip, MPH; Stephanie Rettig, MPH; Marie Samanovic-Golden, PhD; Madalyn Saporito; Pamela Suman; Meron Tasissa; Michael Tuen; Julia Wagner, MPH; James Wilson; Doris Wong, PharmD; Samantha Yip, RN; Heekoung Youn, RN; Lisa Zhao

**Zuckerberg San Francisco General, University of California San Francisco, San Francisco, CA**

Anne F. Luetkemeyer, MD; Chloe Harris, BA; Azquena Munoz Lopez, BS; Daniel Berrner; Dennis Dentoni-Lasofsky, MSN; John Dwyer, RN; Suzanne Hendler, BSN; Elvira Gomez, MPH; WeyLing Phuah, PharmD; Jaime Velasco, BA; Veronica Viar, MS

**Morehouse School of Medicine, Atlanta, GA**

Lilly C. Immergluck, MD; Erica Johnson, PhD; Austin Chan, MD; Fatima Ali, MPH; Sonja Jackson; Noor Mohamed, PharmD; LaKesha Tables, MD; Norberto Fas, MD; Kay Woodson, PharmD; Saadia Khizer, MD; Jacquelyn Ali, MSA; Abdullah Warsama; Eric Gaines; Sierra Jordan Thompson; Cristina Wilson; Trisha Parker, MPH; Xiting Lin; LaTeshia Thomas Seaton, APRN, Derrick Wilson

**Washington University School of Medicine, St. Louis, MO**Rachel M. Presti, MD, PhD; Jane A. O’Halloran, MD, PhD; Ryley M. Thompson; Alem Haile; Kim Gray, NP; Chapelle Ayres; Delaney Carani, RN; Michael Royal; John Tran; Laura Blair; Anita Afghanzada; Natalie Schodl

**NYU VTEU Long Island Research Clinic at NYU Long Island School of Medicine, Mineola, NY**

Martín Bäcker, MD, Kimberly Byrnes, RN, Asif Noor, MD, Steven E. Carsons, MD, Diana Badillo, MD, Sigridh A. Muñoz-Gómez, MD, Andrew B. Fleming, MD, Sajumon K. Joseph, FNP, Sarah J. Pastolero, RN, Maung Aung, Louis Ragolia, PhD, Alicia Vasile, RPh, April Correll, RPh, Christopher Hall, Thomas Palaia, Fiona T. Fitzgerald, Claudia P. De La Matta Rodriguez, MD, Urmee Saha, MD, Miloni H Thakker, MD, Kavita R. Kumar, MD, Lavern Harvey, Lisa Zhao.

**University of Iowa College of Medicine, Iowa City, IA**

Patricia L. Winokur, MD; Jeffery Meier, MD; Jack Stapleton, MD; Laura Stulken, PA; Theresa Hegmann, PA; Deb Pfab, RN; Elizabeth Morgan, RN; Susan Herman, RN; Angel Peguero, CMA; Michelle Rodenburg; Alfred J. Carr; Delilah Johnson

**Howard University College of Medicine, Howard University Hospital, Washington D.C.**

Siham M. Mahgoub, DM; Celia Maxwell, MD; Sarah Shami, PharmD; Edward Bauer, BS; Yuanxiu Chen, MD, PhD; Megan Ware-Pressley, MHA; Debra Ordor, RN; Linda Fletcher, RN; Emmanuel Baidoo, BS; David Jaspan, RPh, MBA; Adetokunbo Adedokun, PharmD, MPH, BCPS; Michelle Strobeck, BS; Michael A. Riga

**University of Alabama at Birmingham, Birmingham, AL**

Paul A. Goepfert, MD; Jenna Weber, RN; Savannah Spaulding, RN; Heather Logan, CRNP; Faye Heard; Foreamben Patel; Michelle Chambers

**Tulane University School of Medicine, New Orleans, LA**

Dahlene N. Fusco, MD; Arnaud C. Drouin, MD; Florice K. Numbi, MD; Hamada F. Rady, PhD; Crystal A. Ward, MSN; Quinn M. Powers, MS; William E. Casey, BS; Brian P. Logarbo, MD; Shae P. Williams, BS; Emily Callegari, MSN

**IDCRC Laboratory Operations Unit – Fred Hutchinson Cancer Center, Seattle, WA**

Christine M. Posavad, PhD; Megan A. Meagher, BS; Michael Stirewalt, MBA; John Hural, PhD; Weston Lawler, BA; Lexi Morritt, MA

**Clinical Monitoring Research Program Directorate, Frederick National Laboratory for Cancer Research, Frederick, MD**

Teri C. Lewis, BS; Lisa A. Giebeig, MS; Theresa M. Engel, MFS.; Caleb J. Griffith, MPH; Wendi L. McDonald, BSN; Alissa E. Burkey, MS; Lisa B. Hoopengardner, MS; Jessica E. Linton, MS; Nikki L. Gettinger, MPH;  Aroussiak Bowen; Beth R. Baseler, MS; Vanessa S. Eccard-Koons, MS; Charles W. R. Hofsommer, JD; Thomas C. Sova, JD; Gary A. Krauss

**Division of Microbiology and Infectious Diseases, National Institute of Allergy and Infectious Diseases, National Institutes of Health, Bethesda, MD**.

Marina Lee, PhD; Mamodikoe Makhene, MD; Mohamed Elsafy, MD; Rhonda Pikaart-Tautges, BS; Janice Arega, MS: Binh Hoang, RPh; Dan Curtin; Hyung Koo, BSN; Elisa Sindall, BSN; Aya Nakamura, RN, MS; Audria Crowder, BS; Guinevere Chun, RN, BSN, MSHS; Frank Kenny, PhD MPH; Seemi Patel, RHP, PharmD; Sonia Gales, MS; Ahsen Khan, JD; Walla Dempsey, PhD; Robert Jurao- RN, BSN; Sonja Crandon, BSN; Seema U. Nayak, MD; Marciela M DeGrace, PhD; Diane J Post, PhD; Paul C Roberts, PhD, John H Beigel, MD SAVE Program

**Supplemental Methods**

Enrollment and Randomization

After informed consent, participants underwent screening, including confirmation of COVID-19 vaccination, medical history, a targeted physical examination, and a urine pregnancy test (if indicated).

Breakthrough SARS-CoV-2 infections

Although the study was not designed to evaluate booster vaccine effectiveness, we collected information on antigen or PCR confirmed symptomatic or asymptomatic SARS-CoV-2 infection at any time after randomization. A nasal swab sample was collected for viral sequencing.

Antigenic Cartography and Antibody Landscape Methods

The antigenic map published in version 2 of Wilks et al. was used as basis for the map shown here and for antibody landscapes. The position of BA.4^1^/BA.5 was estimated as described in section *Estimation of the BA.4/BA.5 position in antigenic space*.^2^ Antigenic cartography uses antibody neutralization data to position virus variants and sera relative to each other in an n-dimensional Euclidean space, in this case a 2-dimensional space, as described by Smith et al^3^ and Wilks et al.^1^ In summary here, for each serum-variant pair, the fold-change from the maximum titer variant in the specific serum is calculated to obtain a target distance from the serum. In an antigenic map, serum and variant coordinates are optimized such that the sum of squared errors between Euclidean map distance and this target distance is minimized, where one distance unit in the map corresponds to one two-fold dilution in the neutralization assay. A variant is positioned by multiple sera and the sera are positioned by their distance to the variant, allowing different locations for individual sera based on the variation in their neutralization profile but each serum contributing to the positioning of a virus variant. For titers below the limit of detection a penalty is introduced in case the serum-variant distance is smaller than target distance, but no penalty is applied to map distances exceeding the target distance. Antibody landscapes^2^ for each study arm and sample day were constructed using the ablandscape.fit function of the ablandscape package^4^ 1.1.0 with the parameters method = "cone", error.sd = 1, bandwidth = 1, degree = 1, control = list (optimise.cone.slope = TRUE). Variant coordinates from the base map were used to fit a single-cone surface to neutralization titers against D614G, Beta, Delta and BA.1 for each serum. Per arm and visit, the surface slope was optimized to match neutralization titers. Samples from non-responding participants, defined as a titer of 20 against D614G, Delta, Beta and BA.1, were removed for landscape making (n = 7 in the uninfected cohort, n=1 in the infected cohort). Surfaces for the subset of samples (n= 87) titrated against BA.4/BA.5 and BA.2.12.1 by the Duke laboratory were constructed the same way with the additional variant titers not considered in the optimization.

A variant is positioned by multiple sera and the sera are positioned by their distance to the variant, allowing different locations for individual sera based on the variation in their neutralization profile but each serum contributing to the positioning of a virus variant.

*Magnitude adjustment of BA.4/BA.5 and BA.2.12.1 titers*

A subset of uninfected Day 15 samples (n=87) was titrated by the Duke laboratory against D614G, BA.1, BA.4/BA.5 and BA.2.12.1. The same samples were titrated by Monogram against D614G, Beta, Delta and BA.1. BA.1 titers corresponded well between both laboratories but Monogram reported higher D614G titers, in line with a previous study.^5^ To match Omicron variant titers to Monogram titrations, data from the Duke laboratory were magnitude adjusted: for each sample, the fold drop from BA.1 to BA.4/BA.5 and BA.2.12.1 in the Duke laboratory was applied to the sample’s BA.1 titer as reported by Monogram.

*Estimation of the BA.4/BA.5 position in antigenic space*

The map in version 2 of Wilks et al.^1^ was used as base map. The antigenic position of BA.4/BA.5 is subject to refinement due to the limited serum groups against which it has currently been titrated. This impacts the positioning of the vertical impulse in Figure 3C, but not the shape of the antibody landscapes as BA.4/BA.5 titers were not used for antibody landscape fitting.

Statistical Analysis

The primary objective of this study is to evaluate the magnitude, breadth and durability of SARS-CoV-2 specific antibody titers in serum samples by estimating 95% confidence intervals (CI) for the geometric mean titer (GMT) at each timepoint when samples are collected. No formal hypothesis tests were planned. Any titer above the lower limit of detection (LLOD) is considered seropositive. 95% CI of GMTs are calculated using the Student’s t-distribution. For the purpose of analysis, participants were defined as previously infected by self-report of a confirmed positive antigen or PCR testing or the detection of anti-SARS-CoV-2 N antibodies.

ANCOVA models were used to estimate GMT ratios of variant vaccines compared to Prototype vaccine and included independent variables for vaccination arms, age (18-64 years and ≥ 65 years of age), previous infection history, and baseline titers. For modeling purposes, titers were log_10_ transformed and estimated mean differences were back transformed to generate GMT ratios between vaccination groups. Unadjusted 97.5% confidence intervals based on the t-distribution are reported.

Study endpoints and sample size determination are included in the protocol.

**TABLE S1: List of COVAIL US Sites**

**Site name**

George Washington University

University of Rochester Medical Center

Hope Clinic of the Emory Vaccine Center

Brigham and Women's Hospital, Harvard Medical School

Saint Louis University

Baylor College of Medicine

University of California San Diego

Emory Center for Childhood Infections and Vaccines of Children’s Healthcare of Atlanta

Duke Human Vaccine Institute, Duke University School of Medicine

University of Illinois at Chicago, Project WISH

University of Texas Medical Branch

Kaiser Permanente Washington Health Research Institute

University of Washington

New York University Manhattan Research Clinic, NYU Grossman School of Medicine

Zuckerberg San Francisco General Hospital, University of California at San Francisco

Morehouse School of Medicine

Washington University School of Medicine

NYU Long Island Research Clinic, NYU Long Island School of Medicine

University of Iowa College of Medicine

Howard University Hospital, Howard University Hospital

University of Alabama at Birmingham

Tulane University School of Medicine

**TABLE S2: Eligible Regimens for Primary Vaccination Series and Boost as of May 2022**

| **Primary series vaccine manufacturer** | **Number of doses in primary series** | **Primary series dose** | **Interval between 1st and 2nd dose** | **Number of booster doses** | **Booster dose** | **Allowed interval between primary series and booster dose*** |
| --- | --- | --- | --- | --- | --- | --- |
| Pfizer BioNTech | 2 | 30 µg | At least 17 days | 1 | 30 µg | ≥ 5 months |
| Moderna | 2 | 100 µg | At least 24 days | 1 | 50 µg | ≥ 5 months |
| Janssen | 1 | 5×10^10^ viral particles | Not applicable | 1 | 5×10^10^ viral particles | ≥ 2 months |

* homologous and heterologous boosters are acceptable

**TABLE S3: Study Arms**

|  | **Arms** | **Vaccine Platform** | **Sample Size** | **Vaccine Candidate** | **Interval**  **(weeks)*** | **Timing of First Dose** | **Timing of Second Dose** |
| --- | --- | --- | --- | --- | --- | --- | --- |
| **Stage 1** | **1** | Moderna mRNA-1273 | 100 | Prototype | ≥16 | D1 | NA |
|  | **2** |  | 100 | Beta (B.1.351) + Omicron BA.1 (B.1.1.529) | ≥16 | D1 | NA |
|  | **3** |  | 100 | Beta (B.1.351) + Omicron BA.1 (B.1.1.529) | ≥16 | D1 | D57 |
|  | **4** |  | 100 | Delta (B.1.617.2) + Omicron BA.1 (B.1.1.529) | ≥16 | D1 | NA |
|  | **5** |  | 100 | Omicron BA.1 (B.1.1.529) | ≥16 | D1 | NA |
|  | **6** |  | 100 | Omicron BA.1 (B.1.1.529) + Prototype | ≥16 | D1 | NA |

*interval (in weeks) since last exposure to SARS-CoV-2 infection or vaccination

**TABLE S4:** **Pseudovirus Neutralization ID_50_ Titers, by Timepoint (baseline and Day 15) and Variant (D614G, Delta (B.1.617.2), Beta (B.1.351) and Omicron BA.1 (B.1.1.529) in All, Uninfected and Infected Participants in all Arms** (Arm 1 or mRNA-1273 [Prototype] vaccine, arms 2 and 3 or Omicron BA.1 + Beta bivalent vaccine, arm 4 or Omicron BA.1 + Delta bivalent vaccine, arm 5 or Omicron BA.1 monovalent vaccine, and arm 6 or Omicron BA.1 + Prototype bivalent vaccine). GMT, geometric mean titer. GMFR, geometric mean fold rise. GMR, geometric mean ratio comparing vaccine arm to mRNA-1273 prototype vaccine. CI, confidence interval

|  | **D614G** | **Delta (B.1.617.2)** | **Beta (B.1.351)** | **Omicron (B.1.1.529)** | **D614G** | **Delta (B.1.617.2)** | **Beta**  **(B.1.351)** | **Omicron (B.1.1.529)** | **D614G** | **Delta (B.1.617.2)** | **Beta (B.1.351)** | **Omicron (B.1.1.529)** |
| --- | --- | --- | --- | --- | --- | --- | --- | --- | --- | --- | --- | --- |
| **Arm 1: mRNA-1273 (Prototype)** | | | | | | | | | | | | |
|  | **Overall (N = 97)** | | | | **Uninfected (N = 76)** | | | | **Infected (N = 21)** | | | |
| **Day 1 GMT^a^**  **(95% CI)** | 3368 (2527, 4489) | 1664 (1218, 2273) | 857  (610, 1204) | 275   (192, 393) | 2313 (1741, 3075) | 1138   (827, 1565) | 550  (395, 766) | 170  (119, 242) | 13117 (7565, 22745) | 6586 (3661, 11848) | 4262 (2117, 8584) | 1573  (812, 3045) |
| **Day 15 GMT**  **(95% CI)** | 20969 (17256, 25480) | 11588  (9428, 14242) | 7690  (6012, 9837) | 2699  (2035, 3578) | 16976 (13851, 20806) | 9327  (7517, 11574) | 5982 (4632, 7724) | 1997  (1482, 2692) | 44587 (30177, 65877) | 25152 (16488, 38369) | 18863 (11081, 32110) | 7906  (4584, 13638) |
| **Day 15 GMFR**  **(95% CI)** | 6.2  (5.2, 7.5) | 7.0   (5.7, 8.6) | 9.1  (7.3, 11.3) | 9.9  (7.7, 12.6) | 7.4  (6.1, 9.0) | 8.3  (6.6, 10.4) | 11.1  (8.8, 14.1) | 11.9  (9.0, 15.9) | 3.4  (2.4, 4.9) | 3.8  (2.7, 5.5) | 4.4  (3.0, 6.4) | 5.0  (3.4, 7.5) |
| **Day 15 seropositive (95% CI)** | 1.00  (0.96, 1.00) | 1.00  (0.96, 1.00) | 1.00  (0.96, 1.00) | 0.98  (0.93, 1.00) | 1.00 (0.95, 1.00) | 1.00  (0.95, 1.00) | 1.00  (0.95, 1.00) | 0.97  (0.91, 1.00) | 1.00  (0.84, 1.00) | 1.00  (0.84, 1.00) | 1.00  (0.84, 1.00) | 1.00  (0.84, 1.00) |
| **Day 15 GMR_D614G_**  **(95.0% CI)** | NA | 0.55  (0.52, 0.58) | 0.37  (0.33, 0.41) | 0.13  (0.11, 0.15) | NA | 0.55  (0.52, 0.58) | 0.35  (0.31, 0.40) | 0.12  (0.10, 0.14) | NA | 0.56   (0.50, 0.64) | 0.42  (0.32, 0.56) | 0.18  (0.12, 0.26) |
| **Arms 2 and 3: mRNA-1273 Beta + Omicron BA.1** | | | | | | | | | | | | |
|  | **Overall (N = 198)** | | | | **Non-Infected (N = 160)** | | | | **Infected (N = 38)** | | | |
| **Day 1 GMT^a^**  **(95% CI)** | 3692  (3020, 4515) | 1685  (1352, 2100) | 967  (765, 1223) | 283  (219, 366) | 2854 (2336, 3487) | 1313  (1061, 1626) | 678  (540, 851) | 195  (152, 251) | 10923 (6562, 18183) | 4819  (2565, 9053) | 4327  (2464, 7597) | 1348  (720, 2524) |
| **Day 15 GMT**  **(95% CI)** | 19760 (16726, 23344) | 11834  (9885, 14167) | 12401 (10326, 14892) | 4784  (3841, 5958) | 16115 (13460, 19292) | 9807  (8197, 11733) | 9603 (7917, 11649) | 3724  (3009, 4608) | 46135 (33789, 62993) | 25847 (15711, 42524) | 35903 (25629, 50295) | 13558  (7262, 25314) |
| **Day 15 GMFR**  **(95% CI)** | 5.3  (4.6, 6.2) | 7.0  (6.0, 8.2) | 12.7  (10.9, 14.9) | 16.8  (13.8, 20.6) | 5.6  (4.9, 6.5) | 7.4  (6.3, 8.8) | 14.1 (12.0, 16.6) | 19.1  (15.6, 23.3) | 4.2  (2.8, 6.4) | 5.4  (3.5, 8.3) | 8.3  (5.3, 12.9) | 10.1  (5.6, 18.1) |
| **Day 15 seropositive (95% CI)** | 0.99  (0.97, 1.00) | 0.99  (0.96, 1.00) | 0.99  (0.97, 1.00) | 0.98  (0.95, 0.99) | 0.99 (0.97, 1.00) | 0.99  (0.97, 1.00) | 0.99 (0.97, 1.00) | 0.99  (0.96, 1.00) | 1.00  (0.91, 1.00) | 0.97  (0.86, 1.00) | 1.00  (0.91, 1.00) | 0.95  (0.82, 0.99) |
| **Day 15 GMR_D614G_**  **(95.0% CI)** | NA | 0.60   (0.55, 0.65) | 0.63  (0.59, 0.67) | 0.24   (0.21, 0.28) | NA | 0.61  (0.58, 0.64) | 0.60  (0.55, 0.64) | 0.23  (0.21, 0.26) | NA | 0.56  (0.38, 0.83) | 0.78  (0.70, 0.87) | 0.29  (0.17, 0.50) |
| **Day 15 GMR_prototype_^b^ (97.5% CI)** | 0.91  (0.72, 1.14) | 1.02  (0.8,1.3) | 1.5  (1.16, 1.95) | 1.75  (1.29, 2.37) | 0.85  (0.66, 1.1) | 0.97   (0.74, 1.27) | 1.4  (1.04, 1.88) | 1.71  (1.23, 2.39) | NA | NA | NA | NA |
| **Arm 4: mRNA-1273 Delta + Omicron** **BA.1** | | | | | | | | | | | | |
|  | **Overall (N = 99)** | | | | **Non-Infected (N = 83)** | | | | **Infected (N = 16)** | | | |
| **Day 1 GMT^a^**  **(95% CI)** | 3519  (2594, 4775) | 1762  (1294, 2400) | 859  (603, 1224) | 273  (190, 393) | 2773 (1997, 3851) | 1379  (988, 1924) | 659  (453, 959) | 208  (143, 303) | 12116 (7145, 20545) | 6290  (3791, 10437) | 3393  (1575, 7310) | 1117  (439, 2839) |
| **Day 15 GMT**  **(95% CI)** | 24337 (19002, 31169) | 14536 (11239, 18801) | 9874  (7311, 13336) | 4503  (3287, 6168) | 21972 (16761, 28802) | 12773  (9703, 16813) | 8608 (6196, 11959) | 3808 (2716, 5337) | 42845 (23664, 77574) | 29734 (15156, 58334) | 21095 (10542, 42212) | 11385 (5224, 24813) |
| **Day 15 GMFR**  **(95% CI)** | 7.1  (5.4, 9.2) | 8.4  (6.4, 11.0) | 11.9  (9.0, 15.7) | 17.1  (12.7, 23.0) | 7.9   (5.8, 10.7) | 9.3  (6.8, 12.7) | 13.1  (9.5, 18.0) | 18.3  (13.1, 25.6) | 3.7  (2.4, 5.6) | 4.8  (3.1, 7.2) | 7.1  (4.5, 11.2) | 11.8  (6.6, 21.1) |
| **Day 15 seropositive (95% CI)** | 0.99  (0.94, 1.00) | 0.99  (0.94, 1.00) | 0.99  (0.94, 1.00) | 0.98  (0.93, 1.00) | 0.99 (0.93, 1.00) | 0.99  (0.93, 1.00) | 0.99  (0.93, 1.00) | 0.98  (0.92, 1.00) | 1.00 (0.78, 1.00) | 1.00  (0.78, 1.00) | 1.00  (0.78, 1.00) | 1.00  (0.78, 1.00) |
| **Day 15 GMR_D614G_**  **(95.0% CI)** | NA | 0.60  (0.56, 0.64) | 0.41  (0.36, 0.46) | 0.19  (0.15, 0.22) | NA | 0.58  (0.55, 0.62) | 0.39  (0.34, 0.45) | 0.17  (0.14, 0.21) | NA | 0.69  (0.57, 0.84) | 0.49  (0.37, 0.65) | 0.27  (0.18, 0.39) |
| **Day 15 GMR_prototype_^b^ (97.5% CI)** | 1.17  (0.9,1.52) | 1.25   (0.94, 1.65) | 1.32   (0.98, 1.77) | 1.73  (1.22, 2.46) | 1.18  (0.88, 1.59) | 1.24   (0.91, 1.69) | 1.28  (0.92, 1.8) | 1.7   (1.16, 2.48) | NA | NA | NA | NA |
| **Arm 5: mRNA-1273 Omicron** **BA.1** | | | | | | | | | | | | |
|  | **Overall (N = 96)** | | | | **Non-Infected (N = 73)** | | | | **Infected (N = 23)** | | | |
| **Day 1 GMT^a^**  **(95% CI)** | 3935  (2953, 5244) | 1946  (1457, 2600) | 956  (658, 1388) | 354   (244, 515) | 2876 (2078, 3980) | 1409  (1020, 1947) | 590  (392, 889) | 210  (141, 312) | 10645 (6972, 16252) | 5421  (3448, 8525) | 4416  (2690, 7249) | 1860  (1086, 3186) |
| **Day 15 GMT**  **(95% CI)** | 19167 (15858, 23168) | 11493  (9408, 14041) | 11161 (8519, 14621) | 6011  (4650, 7771) | 16467 (13087, 20719) | 9490  (7490, 12023) | 8642 (6240, 11968) | 4561  (3379, 6157) | 31039 (24607, 39154) | 21107 (16386, 27189) | 25132 (19186, 32921) | 14441 (10744, 19411) |
| **Day 15 GMFR**  **(95% CI)** | 4.9  (3.9, 6.1) | 5.9  (4.7, 7.4) | 11.7  (9.1, 15.0) | 17.0  (12.9, 22.3) | 5.7  (4.5, 7.4) | 6.7  (5.2, 8.7) | 14.6  (11.1, 19.3) | 21.7  (16.2, 29.0) | 2.9  (1.8, 4.6) | 3.9  (2.4, 6.3) | 5.7  (3.4, 9.5) | 7.8  (4.3, 14.0) |
| **Day 15 seropositive (95% CI)** | 1.00  (0.96, 1.00) | 1.00  (0.96, 1.00) | 0.99  (0.94, 1.00) | 0.99   (0.94, 1.00) | 1.00 (0.95, 1.00) | 1.00  (0.95, 1.00) | 0.99  (0.93, 1.00) | 0.99  (0.93, 1.00) | 1.00  (0.85, 1.00) | 1.00  (0.85, 1.00) | 1.00  (0.85, 1.00) | 1.00  (0.85, 1.00) |
| **Day 15 GMR_D614G_**  **(95.0% CI)** | NA | 0.60  (0.57, 0.64) | 0.58  (0.50, 0.68) | 0.31  (0.27, 0.37) | NA | 0.58  (0.54, 0.62) | 0.52  (0.43, 0.64) | 0.28  (0.23, 0.34) | NA | 0.68  (0.61, 0.76) | 0.81  (0.71, 0.93) | 0.47  (0.39, 0.56) |
| **Day 15 GMR_prototype_^b^ (97.5% CI)** | 0.85  (0.65,1.1) | 0.91  (0.69, 1.21) | 1.36   (1, 1.83) | 1.94   (1.36, 2.76) | 0.87  (0.64, 1.18) | 0.91  (0.66, 1.26) | 1.38  (0.97, 1.95) | 2.03  (1.37, 3) | NA | NA | NA | NA |
| **Arm 6: mRNA-1273 Omicron BA.1 + Prototype** | | | | | | | | | | | | |
|  | **Overall (N = 96)** | | | | **Non-Infected (N = 76)** | | | | **Infected (N = 20)** | | | |
| **Day 1 GMT^a^**  **(95% CI)** | 3503  (2521, 4867) | 1738  (1250, 2417) | 958  (676, 1359) | 352  (244, 508) | 2598 (1889, 3574) | 1292  (929, 1797) | 651  (468, 905) | 230   (164, 324) | 10902 (4390, 27076) | 5370  (2290, 12596) | 4165  (1674, 10361) | 1763  (673, 4619) |
| **Day 15 GMT**  **(95% CI)** | 26125 (21593, 31607) | 13809 (10857, 17564) | 10742 (8164, 14135) | 5322  (3986, 7106) | 21096 (17398, 25580) | 10798  (8331, 13995) | 7814 (5879, 10386) | 3747  (2773, 5061) | 57011 (38113, 85280) | 33893 (22070, 52050) | 34324 (20763, 56743) | 19160 (11875, 30914) |
| **Day 15 GMFR**  **(95% CI)** | 7.2  (5.5, 9.4) | 7.7  (5.9, 9.9) | 10.9  (8.4, 14.1) | 14.6  (11.0, 19.5) | 7.8  (6.0, 10.2) | 8.1  (6.2, 10.5) | 11.7  (9.1, 15.1) | 15.8  (12.1, 20.8) | 5.2  (2.3, 12.1) | 6.3  (2.8, 14.1) | 8.2  (3.5, 19.4) | 10.9  (4.1, 28.6) |
| **Day 15 seropositive (95% CI)** | 1.00  (0.96, 1.00) | 0.99  (0.94, 1.00) | 0.99  (0.94, 1.00) | 0.99  (0.94, 1.00) | 1.00 (0.95, 1.00) | 0.99  (0.93, 1.00) | 0.99  (0.93, 1.00) | 0.99   (0.93, 1.00) | 1.00  (0.83, 1.00) | 1.00  (0.83, 1.00) | 1.00  (0.83, 1.00) | 1.00  (0.83, 1.00) |
| **Day 15 GMR_D614G_**  **(95.0% CI)** | NA | 0.53  (0.45, 0.62) | 0.41  (0.35, 0.49) | 0.20  (0.17, 0.24) | NA | 0.51  (0.42, 0.62) | 0.37  (0.30, 0.45) | 0.18  (0.14, 0.22) | NA | 0.59   (0.50, 0.71) | 0.60   (0.48, 0.75) | 0.34   (0.27, 0.41) |
| **Day 15 GMR_prototype_^b^ (97.5% CI)** | 1.2  (0.92, 1.57) | 1.14   (0.86, 1.52) | 1.29  (0.95, 1.74) | 1.71  (1.19, 2.43) | 1.15  (0.85, 1.56) | 1.06  (0.77, 1.46) | 1.16  (0.82, 1.64) | 1.56  (1.06, 2.31) | NA | NA | NA | NA |

N = Number of subjects with results available at time point. Eleven subjects infected after enrollment and before Day 15 were excluded from the analysis.  
Confidence intervals of the geometric means were calculated with the Student’s t distribution on log-transformed data

^a^ Day 1 pre-boost GMT

^b^ Based on ANCOVA modeling and comparison to Prototype vaccine; the model includes adjustment for pre-booster antibody titers, infection history and age. Confidence intervals are unadjusted at level 97.5%.

**TABLE S5:** **Pseudovirus Neutralization ID_50_ Titers, by Timepoint (baseline and Day 15) and Variant (D614G, Omicron (B.1.1.529, BA.2.12.1. and BA.4/BA.5) in All and Uninfected Participants in all Arms** (Arm1 or Prototype monovalent vaccine, arms 2 and 3 or Omicron BA.1 + Beta bivalent vaccine, arm 4 or Omicron BA.1 + Delta bivalent vaccine, arm 6 or Omicron BA.1 + Prototype bivalent vaccine). GMT, geometric mean titer. GMFR, geometric mean fold rise. GMR, geometric mean ratio to D614G. CI, confidence interval

|  | **D614G** | **Omicron (B.1.1.529)** | **Omicron**  **(BA.2.12.1)** | **Omicron (BA.4/BA.5)** | **D614G** | **Omicron (B.1.1.529)** | **Omicron**  **(BA.2.12.1)** | **Omicron (BA.4/BA.5)** |
| --- | --- | --- | --- | --- | --- | --- | --- | --- |
| **Arm 1: mRNA-1273 (Prototype)** | | | | | | | | |
|  | **Overall (N = 23)** | | | | **Uninfected (N = 20)** | | | |
| **Day 15 GMT**  **(95% CI)** | 3580  (2147, 5969) | 1808  (1115, 2932) | 1179  (726, 1917) | 628  (387, 1019) | 3320  (1887, 5843) | 1503  (949, 2381) | 936  (632, 1387) | 517 (324, 826) |
| **Day 15 seropositive**  **(95% CI)** | 1.00  (0.85, 1.00) | 1.00  (0.85, 1.00) | 1.00  (0.85, 1.00) | 1.00  (0.85, 1.00) | 1.00  (0.83, 1.00) | 1.00  (0.83, 1.00) | 1.00  (0.83, 1.00) | 1.00 (0.83, 1.00) |
| **Day 15 GMR_D614G_**  **(95.0% CI)** | NA | 0.51  (0.35, 0.72) | 0.33  (0.23, 0.48) | 0.18  (0.13, 0.24) | NA | 0.45  (0.31, 0.66) | 0.28  (0.20, 0.41) | 0.16 (0.11, 0.21) |
| **Arm 2: Beta + Omicron BA.1** | | | | | | | | |
|  | **Overall (N = 23)** | | | | **Uninfected (N = 21)** | | | |
| **Day 15 GMT**  **(95% CI)** | 3748  (2274, 6178) | 2713  1377, 5345) | 1579  (805, 3097) | 784  (438, 1403) | 2993 (1969, 4551) | 2125 (1139, 3965) | 1279 (661, 2471) | 628 (367, 1074) |
| **Day 15 seropositive**  **(95% CI)** | 1.00  (0.85, 1.00) | 0.96  (0.78, 1.00) | 0.96  (0.78, 1.00) | 1.00  (0.85, 1.00) | 1.00 (0.84, 1.00) | 0.95 (0.76, 1.00) | 0.95 (0.76, 1.00) | 1.00 (0.84, 1.00) |
| **Day 15 GMR_D614G_**  **(95.0% CI)** | NA | 0.72  (0.50, 1.05) | 0.42  (0.29, 0.60) | 0.21  (0.16, 0.27) | NA | 0.71 (0.48, 1.06) | 0.43 (0.29, 0.63) | 0.21 (0.16, 0.28) |
| **Arm 4: Delta + Omicron BA.1** | | | | | | | | |
|  | **Overall (N = 25)** | | | | **Uninfected (N = 24)** | | | |
| **Day 15 GMT**  **(95% CI)** | 4807  (3231, 7151) | 2375  (1309, 4310) | 1735  (910, 3306) | 809  (473, 1382) | 4593 (3069, 6876) | 2242 (1218, 4128) | 1660 (852, 3234) | 765 (443, 1322) |
| **Day 15 seropositive**  **(95% CI)** | 1.00  (0.86, 1.00) | 1.00  (0.86, 1.00) | 1.00  (0.86, 1.00) | 1.00  (0.86, 1.00) | 1.00  (0.86, 1.00) | 1.00  (0.86, 1.00) | 1.00  (0.86, 1.00) | 1.00 (0.86, 1.00) |
| **Day 15 GMR_D614G_**  **(95.0% CI)** | NA | 0.49  (0.36, 0.68) | 0.36  (0.26, 0.51) | 0.17  (0.13, 0.22) | NA | 0.49  (0.35, 0.68) | 0.36  (0.25, 0.52) | 0.17 (0.13, 0.22) |
| **Arm 6: Omicron BA.1 + Prototype** | | | | | | | | |
|  | **Overall (N = 24^a^)** | | | | **Uninfected (N = 22^b^)** | | | |
| **Day 15 GMT**  **(95% CI)** | 4484  (2954, 6809) | 2507  (1525, 4122) | 1756  (978, 3151) | 817  (495, 1350) | 3748 (2651, 5298) | 1972 (1337, 2907) | 1338 (834, 2148) | 635 (447, 903) |
| **Day 15 seropositive**  **(95% CI)** | 1.00  (0.86, 1.00) | 1.00  (0.85, 1.00) | 1.00  (0.85, 1.00) | 1.00  (0.86, 1.00) | 1.00 (0.85, 1.00) | 1.00 (0.84, 1.00) | 1.00 (0.84, 1.00) | 1.00 (0.85, 1.00) |
| **Day 15 GMR_D614G_**  **(95.0% CI)** | NA | 0.56  (0.42, 0.73) | 0.39  (0.28, 0.54) | 0.18  (0.14, 0.24) | NA | 0.53 (0.40, 0.71) | 0.36 (0.26, 0.50) | 0.17 (0.13, 0.22) |

^a^N was 24 for D614G and BA.4/BA.5 and 23 for Omicron B.1.1.529 and BA.2.12.1.

^b^N was 22 for D614G and BA.4/BA.5 and 21 for Omicron B.1.1.529 and BA.2.12.1.

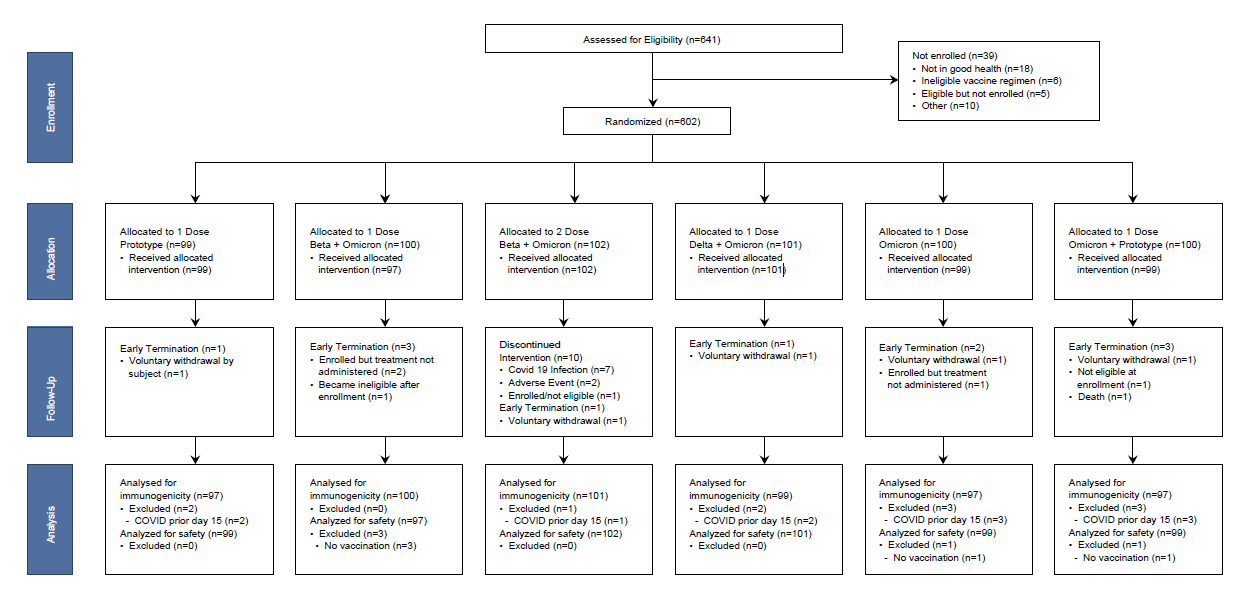
**FIGURE S1: Consort Diagram for Stage 1:** All participants were randomly assigned to a Moderna mRNA vaccine (50 ug total dose) as described in the allocation line of the diagram.

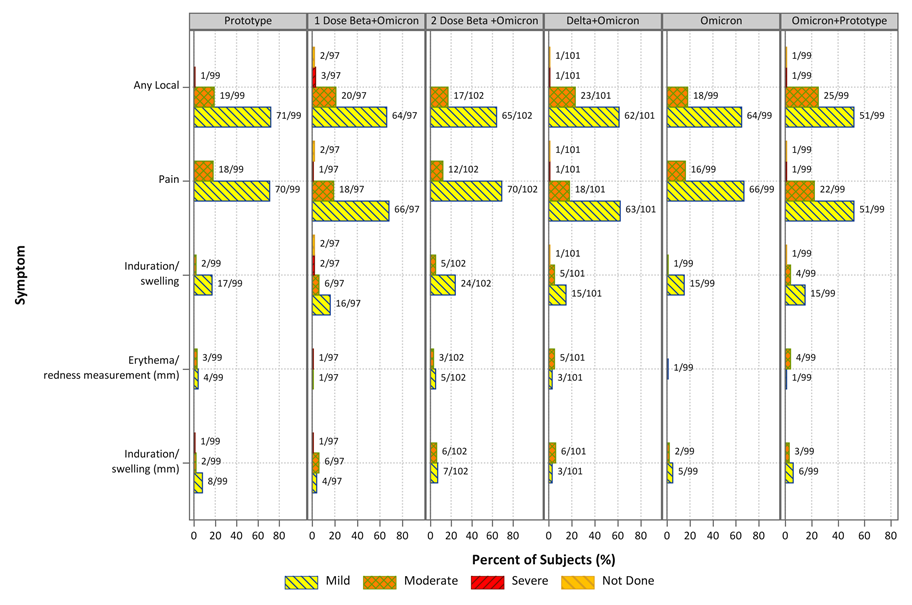
**FIGURE S2: Reactogenicity Reported up to 7 Days after Vaccination**

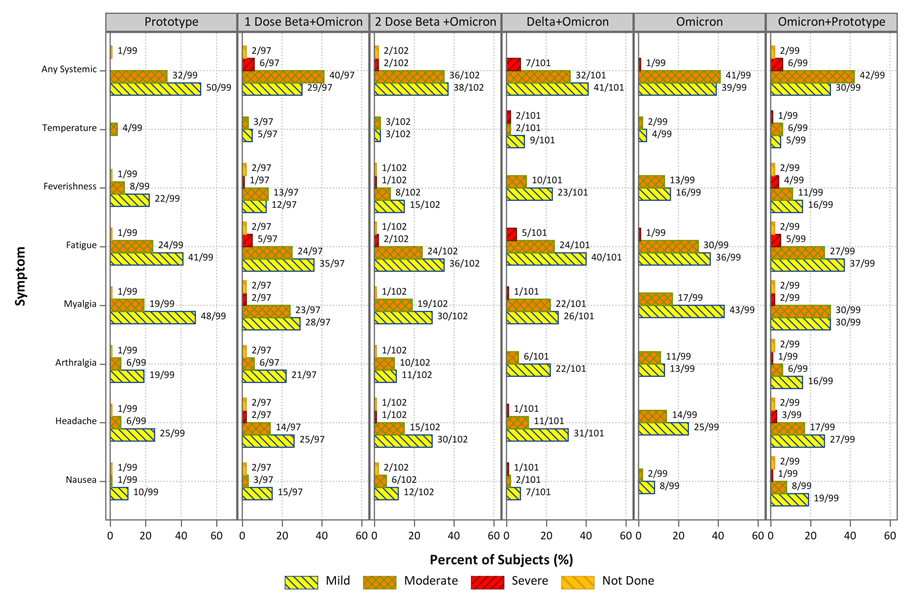

**FIGURE S3: Unsolicited Adverse Events Reported through Day 29 after Vaccination**

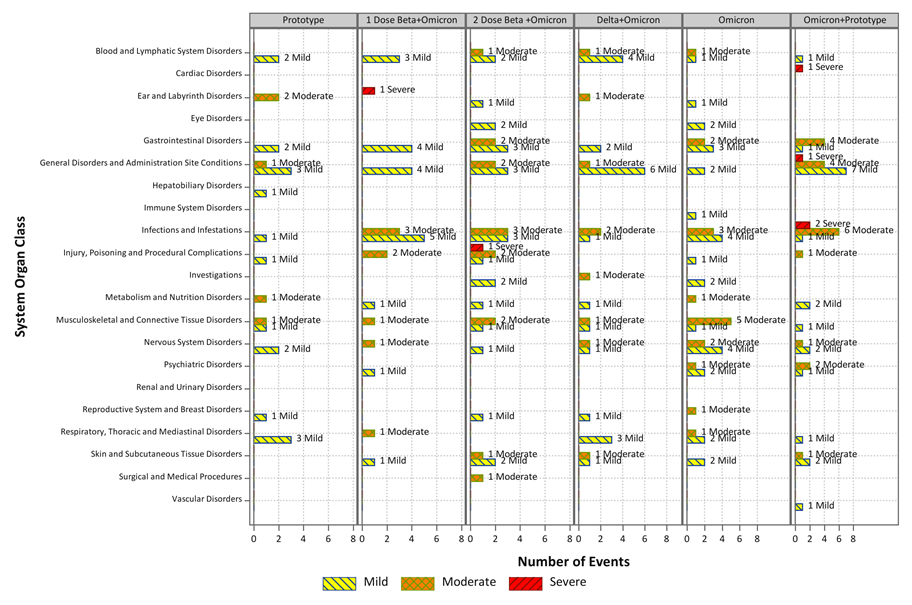
